# Integrative polygenic risk score improves the prediction accuracy of complex traits and diseases

**DOI:** 10.1101/2023.02.21.23286110

**Authors:** Buu Truong, Leland E. Hull, Yunfeng Ruan, Qin Qin Huang, Whitney Hornsby, Hilary Martin, David A. van Heel, Ying Wang, Alicia R. Martin, S. Hong Lee, Pradeep Natarajan

## Abstract

Polygenic risk scores (PRS) are an emerging tool to predict the clinical phenotypes and outcomes of individuals. Validation and transferability of existing PRS across independent datasets and diverse ancestries are limited, which hinders the practical utility and exacerbates health disparities. We propose PRSmix, a framework that evaluates and leverages the PRS corpus of a target trait to improve prediction accuracy, and PRSmix+, which incorporates genetically correlated traits to better capture the human genetic architecture. We applied PRSmix to 47 and 32 diseases/traits in European and South Asian ancestries, respectively. PRSmix demonstrated a mean prediction accuracy improvement of 1.20-fold (95% CI: [1.10; 1.3]; P-value = 9.17 × 10^−5^) and 1.19-fold (95% CI: [1.11; 1.27]; P-value = 1.92 × 10^−6^), and PRSmix+ improved the prediction accuracy by 1.72-fold (95% CI: [1.40; 2.04]; P-value = 7.58 × 10^−6^) and 1.42-fold (95% CI: [1.25; 1.59]; P-value = 8.01 × 10^−7^) in European and South Asian ancestries, respectively. Compared to the previously established cross-trait-combination method with scores from pre-defined correlated traits, we demonstrated that our method can improve prediction accuracy for coronary artery disease up to 3.27-fold (95% CI: [2.1; 4.44]; P-value after FDR correction = 2.6 × 10^−4^). Our method provides a comprehensive framework to benchmark and leverage the combined power of PRS for maximal performance in a desired target population.

## INTRODUCTION

Thousands of polygenic risk scores (PRS) have been developed to predict an individual’s genetic propensity to diverse phenotypes^1^. PRS are generated when risk alleles for distinct phenotypes are weighted by their effect size estimates and summed^2^. Risk alleles included in PRS have traditionally been identified from genome-wide association studies (GWAS) results conducted on a training dataset, which are weighted and aggregated to derive a PRS to predict distinct phenotypes. The association between PRS and the phenotype of interest is subsequently evaluated in a test dataset that is non-overlapping with the training dataset^3^.

Most PRS have been developed in specific cohorts that may vary in terms of population demographics, admixture, environment, and SNP availability. Limited validation of many PRS outside of the training datasets and poor transferability of PRS to other populations may limit their clinical utility. However, pooling of data from individual PRS generated and validated in diverse cohorts has the potential to improve the predictive ability of PRS across diverse populations. The Polygenic Score Catalog (PGS Catalog) is a publicly available repository that archives SNP effect sizes for PRS estimation. The SNP effect sizes were developed from various methods (e.g. P+T^4^, LDpred^5,6^, PRS-CS^7^, etc.) to obtain the highest prediction accuracy in the studied dataset. PRS metadata enables researchers to replicate PRS in independent cohorts and aggregate SNP effects to refine PRS and enhance the accuracy and generalizability in broader populations^8^. However, optimizing PRS performance requires methodological approaches to adjust GWAS estimate effect sizes that take into account correlated SNPs (i.e., linkage disequilibrium) and refine PRS for the target population^4,5,7,9–12^. Furthermore, numerous scores are often present for single traits with varied validation metrics in non-overlapping cohorts. There is a lack of standardized approaches combining PRS from this growing corpus to enhance prediction accuracy and generalizability while minimizing bias, for a target cohort^8,11,13^.

To address these issues, we sought to: 1) validate previously developed PRS in two geographically and ancestrally distinct cohorts, the *All of Us* Research Program (AoU) and the Genes & Health cohort, and 2) present and evaluate new methods for combining previously calculated PRS to maximize performance beyond all best performing published PRS. To better capture the genetic architecture of the outcome traits, we proposed PRSmix, a framework to combine PRS from the same trait with the outcome trait. Previous studies highlighted the effect of pleiotropic information on a trait’s genetic architecture^14,15^. Therefore, we proposed PRSmix+ to additionally combine PRS from other genetically correlated traits to further improve the PRS for a given trait.

To assess the prediction improvement, we performed PRSmix and PRSmix+ for 47 traits in European ancestry and 32 traits in South Asian ancestry. We evaluated 1) the relative improvement of the proposed framework over the best-performing pre-existing PRS for each trait, 2) the efficient training sample sizes required to improve the PRS, 3) the predictive improvement in 6 groups including anthropometrics, blood counts, cancer, cardiometabolic, biochemistry and other conditions as the prediction accuracies varied in each group, and 4) the clinical utility and pleiotropic effect of the newly built PRS for coronary artery disease. Overall, we show that PRSmix and PRSmix+ significantly improved prediction accuracy. An R package for preprocessing and harmonizing the SNP effects from the PGS Catalog as well as assessing and combining the scores was developed to facilitate the combining of pre-existing PRS scores for both ancestry-specific and cross-ancestry contexts using the totality of published PRS. The development of this framework has the potential to improve precision health by improving the generalizability in the application of PRS^16^.

## RESULTS

### Overview of methods

A single PRS may only reflect genetic effects captured in the discovery dataset of a single study that may be only a part of the total genetic effects underlying the trait of interest. Therefore, we harmonized and combined multiple sets of PRS to establish a new set of scores, which gather information across studies and traits. Our approach leveraged multiple well-powered PRSs to improve prediction accuracy and is detailed in Fig. 1.

**Figure 1.**
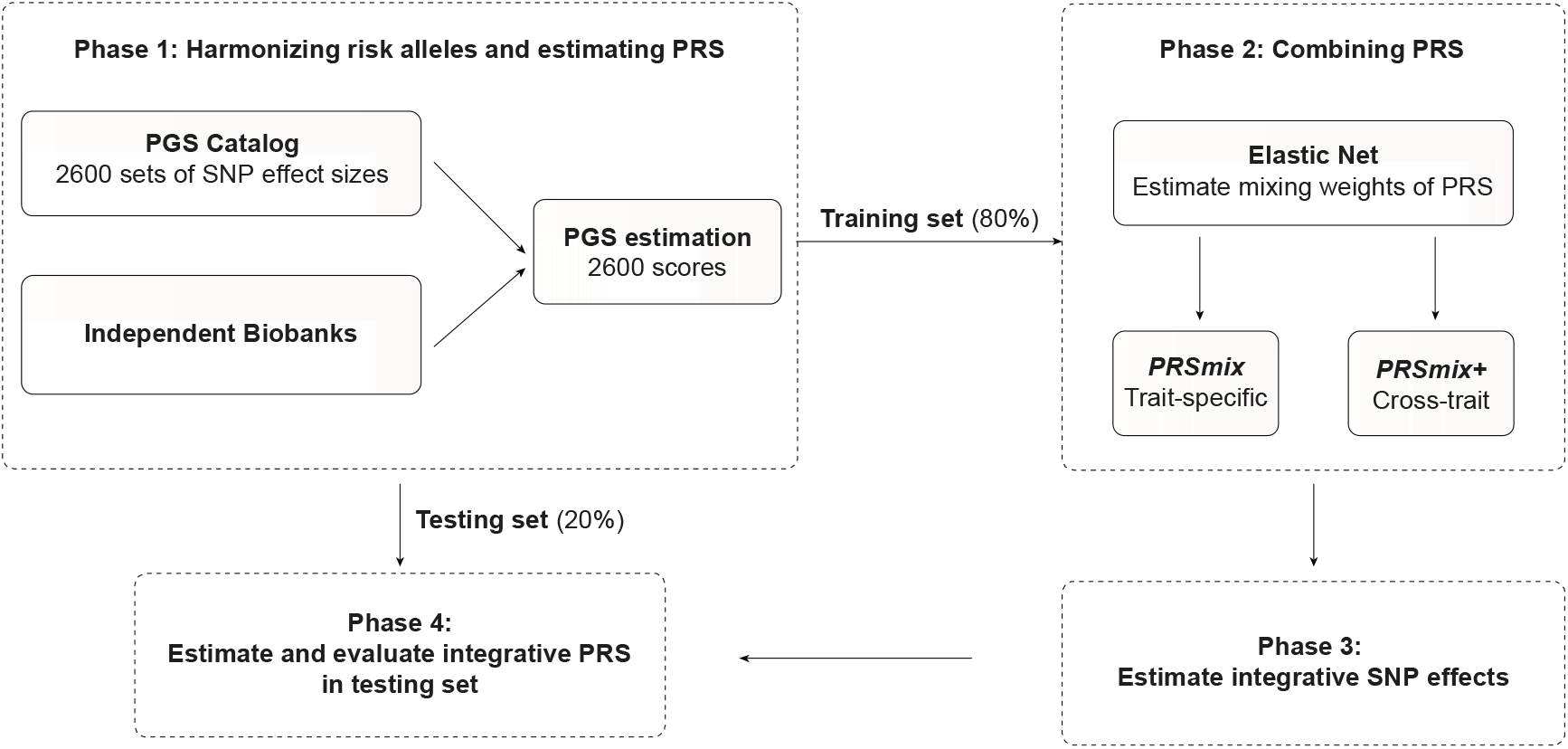
The framework of the trait-specific and cross-trait PRS integration. In Phase 1, we obtained the SNP effects from the PGS Catalog and then harmonized the effect alleles as the alternative alleles in the independent cohorts. In each independent biobank (*All of Us*, Genes & Health), we estimated the PRS and split the data into training (80%) and testing (20%) datasets. In Phase 2, in the training dataset, we trained the Elastic Net model with high-power scores to estimate the mixing weights for the PRSs. The training phase could include PRSs from traits corresponding to outcomes (PRSmix) or all traits (PRSmix+). The training was adjusted for age, sex, and 10 principal components (PCs). In Phase 3, we adjusted the per-allele effect sizes from each single PRS by multiplying with the corresponding mixing weights obtained in the training phase. The final per-allele effect sizes are estimated as the weighted sum of the SNP effects across different single scores. In Phase 4, we evaluated the re-estimated per-allele effect sizes in the testing dataset.

Our combination frameworks leveraged the PGS Catalog^17^ as the resource of SNP effects to estimate single PRSs. To avoid overfitting, we used *All of Us* and Genes & Health cohorts (see Methods) due to non-overlapping samples from the original GWAS. We randomly divided the target cohort into a training set (80%) and a testing set (20%). We selected the most common traits from the PGS Catalog which have the highest number of PRS. For the stability of the linear combination, we curated binary traits with a prevalence > 2% in the target cohort. Continuous traits were assessed using partial R^2^ which is estimated as the difference between the full model of PRS and covariates (age, sex, and 10 PCs) and the null model of only covariates. For binary traits, the prediction accuracy was converted to liability R^2^ with disease prevalence approximated as the prevalence in the corresponding cohort.

To combine the scores, we employed Elastic Net^18^ to construct linear combinations of the PRS. We proposed two combination frameworks: 1) PRSmix combines the scores developed from the same outcome trait, and 2) PRSmix+ combines all the high-power scores across other traits. Trait-specific combinations, PRSmix, can leverage the PRSs developed from different studies and methods to more fully capture the genetic effects underlying the traits. It has also been shown that complex traits are determined by genes with pleiotropic effects^15^. Therefore, we additionally proposed a cross-trait combination, PRSmix+, to make use of pleiotropic effects and further improve prediction accuracy.

First, we evaluated the improvement for each method, defined as the fold-ratio of the method compared to the prediction accuracy of the best single PRS. For a fair comparison with the proposed framework, we selected the best single PRS from the training set and evaluated its performance in the testing set. First, we performed simulations to assess the improvement with various heritabilities and training sample sizes. We estimated the slope of improvement of prediction accuracy by increasing training sample sizes for various heritabilities.

Next, we applied the proposed frameworks in two distinct cohorts; (1) the *All of Us* program, in which 47 traits were tested in U.S. residents of European ancestry, and (2) the Genes & Health (G&H) cohort, in which 32 traits were tested in British South Asian ancestry (Supplementary Table 1). In each cohort, we compared the improvement of our proposed framework with the single best score from the PGS Catalog. We estimated the averaged fold-ratio as a measure of the improvement of prediction accuracy by our approach, compared to the best single score from PGS Catalog. We also classified the traits into 6 categories as anthropometrics, blood counts, cancer, cardiometabolic, biochemistry, and other conditions (Supplementary Table 2 and 3). Cancer traits were not considered in the younger Genes & Health cohort due to their low prevalence (<2%). We then present additional detailed analyses for coronary artery disease focused on clinical utility improvements relative to existing PRS.

### Simulations were used to evaluate the combination frameworks

To compare the performance of PRSmix and PRSmix+ against the best single PRS and evaluate the sample sizes needed for training the mixing weights, we performed simulations with real genotypes of European ancestry in the UK Biobank given the large sample sizes available (Fig. 2). Briefly, we randomly split 7,000 individuals as a testing data set mimicking the testing size of 20% of real data. In the remaining dataset, we used 200,000 individuals for GWAS to estimate the SNP effect sizes for PRS calculations. Finally, with the rest of the data, we randomly selected different sample sizes as the training sample to evaluate the sample sizes needed to train the mixing weights. To assess the improvement of PRS performance, we computed the fold-ratio of prediction accuracy R^2^ between PRSmix and PRSmix+ against the best-performing single simulated PRS.

**Figure 2.**
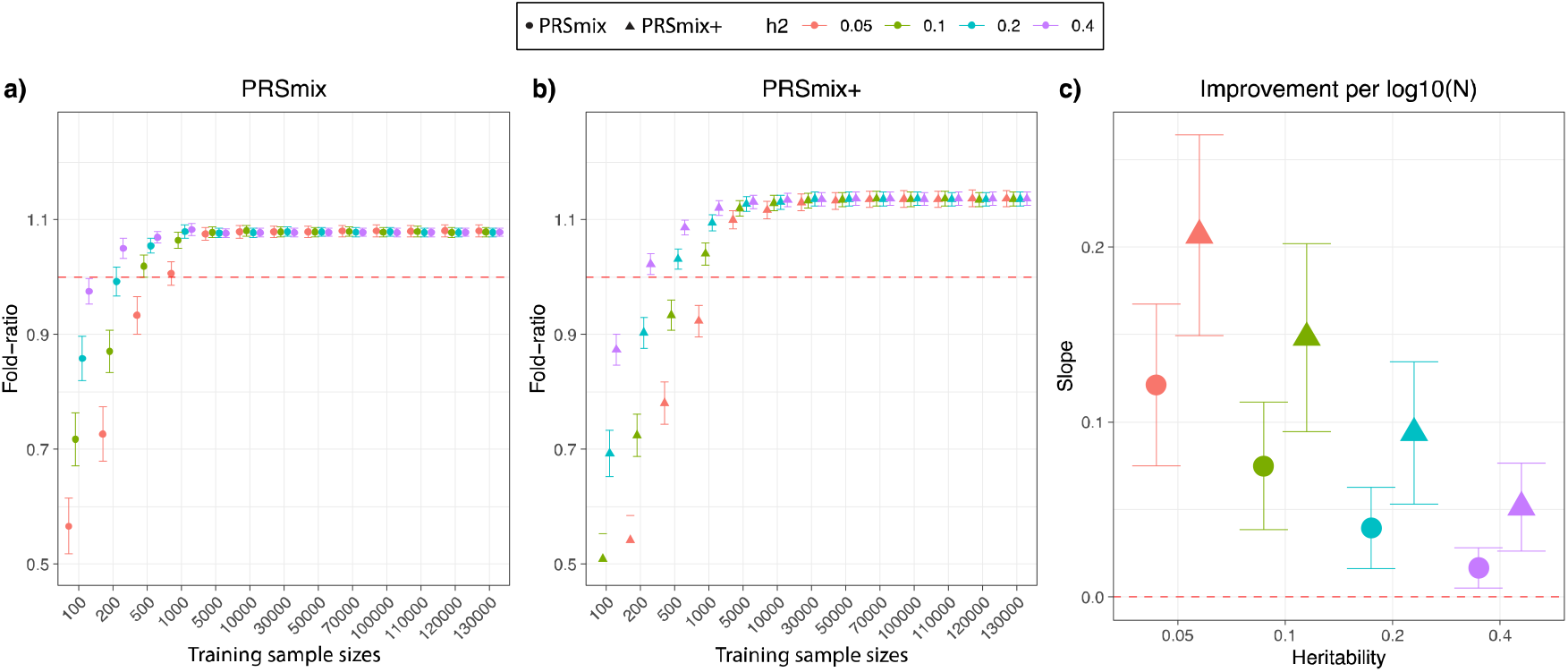
Simulations to demonstrate the predictive improvement of PRSmix and PRSmix+. The points and triangles represent the mean fold-ratio of R^2^ between **(a)** PRSmix and **(b)** PRSmix+, respectively, versus the best single PRS. **(c)** The improvement per logarithm with base 10 of sample size for various heritabilities was represented as a slope of a linear regression of fold-ratio ~ log10(N). In simulations, the correlation within simulated trait-specific PRSs was 0.8, and the correlation between trait-specific and correlated PRSs was 0.4 (see Methods). The whiskers demonstrate confidence intervals across 200 replications. The dashed red lines represent the reference for fold-ratio equal 1 for (a) and (b), and equal 0 for (c).

Our results showed that the trait-specific combination, PRSmix, showed no improvement with the training sample smaller than 500 for most of the traits. Our simulations illustrated that traits with low heritability required a larger sample size to achieve an improvement compared to traits with high heritability (Fig. 2a and 2b). PRSmix demonstrated a better performance compared to the best single PRS with training sample sizes from N_training_ = 200 samples for the high heritable trait (h^2^ = 0.4) to N_training_ = 5000 samples for the low heritable trait (h^2^=0.05) (Fig. 2a and 2b). We observed that PRSmix demonstrated a saturation of improvement from N_training_ = 10,000. PRSmix+ demonstrated negligible further improvement when the training sample size was increased from 30,000 but maintained consistent improvement relative to PRSmix and the best single PRS. Moreover, we observed that traits with higher heritability or higher best prediction accuracy of a single PRS demonstrated a smaller improvement compared to traits with a smaller heritability (Fig. 2c).

### Combining trait-specific PRS improves prediction accuracy (PRSmix)

To determine if a trait-specific combination, namely PRSmix, would improve the accuracy of PRS prediction, we used data from European ancestry participants in the *All of Us* research program who had undergone whole genome sequencing, and Genes & Health participants of South Asian ancestry. We randomly split the independent cohorts into training (80%) and testing sets (20%). The training set was used to train the weights of each PRS, referred as mixing weights, that indicate how much each PRS explain the phenotypic variance in the training set, and the PRS accuracies were evaluated in the testing set (Fig. 1). We curated 47 traits and 32 traits in the *All of Us* and Genes & Health cohorts, respectively. For binary traits, we removed traits with a prevalence of smaller than 2% (see Methods, Supplementary Table 1). Traits with the best-performance trait-specific single PRS which showed a lack of power were also removed. Overall, we observed a significant improvement compared to 1 using a two-tailed paired t-test with PRSmix. PRSmix significantly improves the prediction accuracy compared to the best PRS estimated from the PGS Catalog. PRSmix improved 1.20-fold (95% CI: [1.10; 1.3]; P-value = 9.17 × 10^−5^) and 1.19-fold (95% CI: [1.11; 1.27]; P-value = 1.92 × 10^−6^) compared to the best PRS from PGS Catalog for European ancestry and South Asian ancestry, respectively.

In European ancestry, we observed the greatest improvement of PRSmix against the best single PRS for rheumatoid arthritis of 3.36-fold. Furthermore, in South Asian ancestry, we observed that PRSmix of coronary artery disease had the best improvement of 2.32-fold compared to the best-performance single PRS. Details of the prediction accuracy are shown in Supplementary Fig. 1, 2 and Supplementary Table 2, 3. This was consistent with findings in simulations since traits with a lower single PRS performance demonstrated a better improvement with the combination strategy.

### Cross-trait combination further improved PRS accuracy and highlighted the contribution of pleiotropic effects (PRSmix+)

We next assessed the contribution of pleiotropic effects from cross-trait PRSs to determine if these would further improve the combination framework (PRSmix+), by including high-power PRSs from within 2600 PRSs in the PGS Catalog. To evaluate the power of PRS and improve computational efficiency, we employed the theoretic power and variance of partial R^2^ for continuous traits and liability R^2^ for binary traits (see Methods). We observed that PRSmix+ further improved the prediction accuracy compared to the best PGS Catalog in European ancestry (Fig. 3a) and South Asian ancestry (Fig. 3b). We observed an improvement of 1.72-fold (95% CI: [1.40; 2.04]; P-value = 7.58 × 10^−6^) and 1.42-fold (95% CI: [1.25; 1.59]; P-value = 8.01 × 10^−7^) higher compared to the best PGS Catalog for European ancestry and South Asian ancestry, respectively. PRSmix+ significantly improved the prediction accuracy compared to PRSmix, in both European and South Asian ancestry with 1.46-fold (95% CI: [1.17; 1.75]; P-value = 0.002) and 1.19-fold (95% CI: [1.07; 1.32]; P-value = 0.001), respectively (Supplementary Fig. 3).

**Figure 3.**
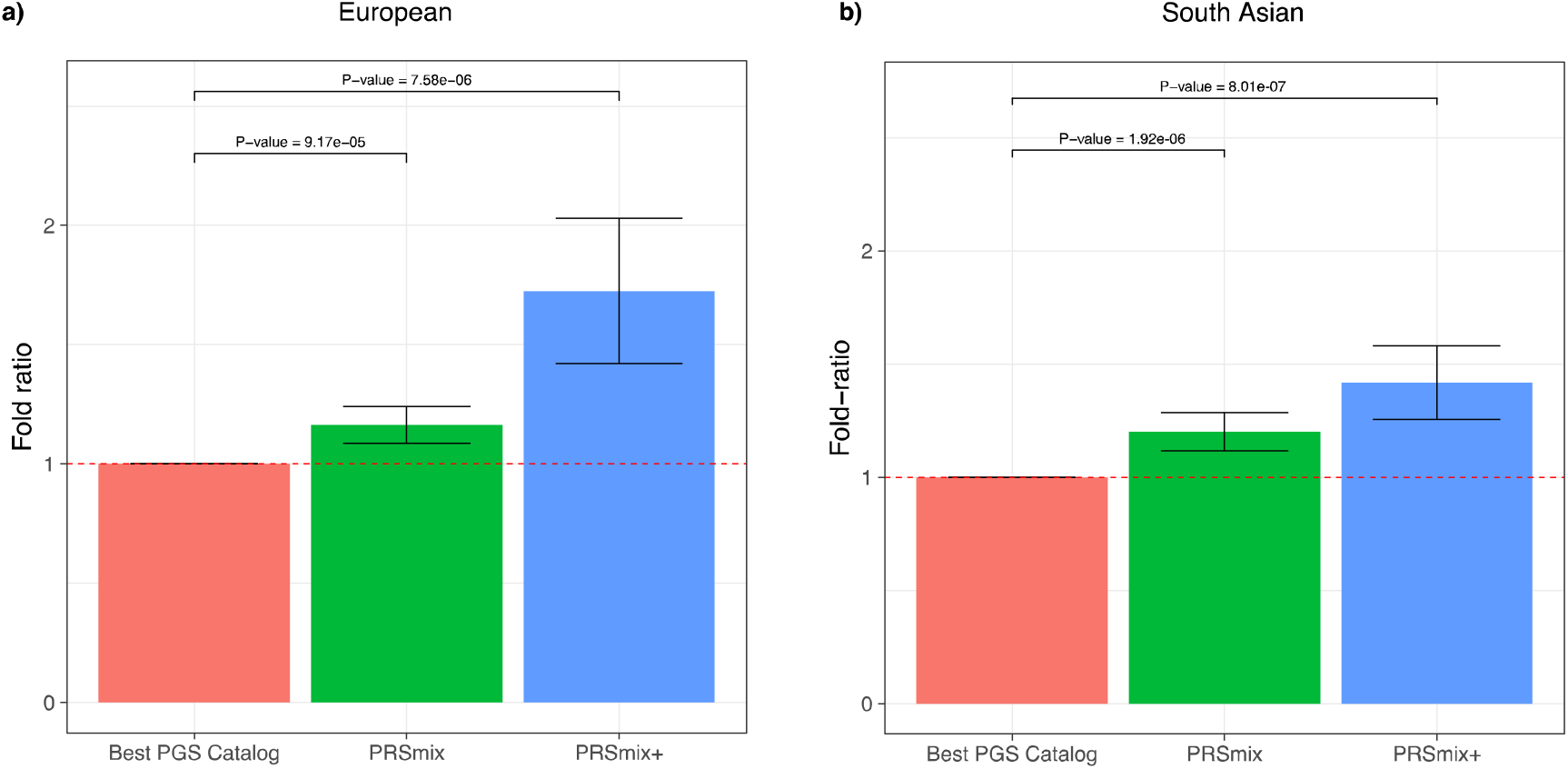
Comparison of PRSmix and PRSmix+ versus the best PGS Catalog in European and South Asian ancestries. The relative improvement compared to the best single PRS was assessed in **(a)** the European ancestry in the *All of US* cohort and **(b)** South Asian ancestry in the Genes & Health cohort. PRSmix combines trait-specific PRSs and PRSmix+ combines additional PRSs from other traits. The best PGS Catalog score was selected by the best performance trait-specific score in the training sample and evaluated in the testing sample. The prediction accuracy (R^2^) was calculated as partial R^2^ which is a difference of R^2^ between the model with PRS and covariates including age, sex, and 10 PCs versus the base model with only covariates. Prediction accuracy for binary traits was assessed with liability-R^2^ where disease prevalence was approximately estimated as a proportion of cases in the testing set. The whiskers reflect the maximum and minimum values within the 1.5 × interquartile range. The bars represent the ratio of prediction accuracy of PRSmix and PRSmix+ versus the best PRS from the PGS Catalog across 47 traits and 32 traits in *All of Us* and Genes and Heath cohorts, respectively, and the whiskers demonstrate 95% confidence intervals. P-values for significance difference of the fold-ratio from 1 using a two-tailed paired t-test. PRS: Polygenic risk scores.

Consistent with our simulation results, a smaller improvement was observed for traits with a higher baseline prediction accuracy from PGS Catalog (Supplementary Fig. 4), noting that the baseline prediction accuracy depends on the heritability and genetic architecture (i.e. polygenicity). In contrast, more improvement was observed for traits with lower heritability, thus lower prediction accuracy, when comparing the single best PRS (Fig. 1c).

### Prediction accuracy and predictive improvement across various types of traits

We next compared PRSmix and PRSmix+ with the best PRS estimated from the PGS Catalog across 6 categories, including anthropometrics, blood counts, cancer, cardiometabolic, biochemistry, and other conditions (see Methods). PRSmix demonstrates a higher prediction accuracy across all types of traits in both European and South Asian ancestries (Fig. 4). We observed a similar trend in the predictive performance of PRSmix+ across different types of traits. In European, the smallest improvement with PRSmix+ was in anthropometric traits of 1.14-fold (95% CI: [1.03; 1.25]; P-value = 0.01) while “other conditions” (including depression, asthma, migraine, current smoker, hypothyroid, osteoporosis, glaucoma, rheumatoid arthritis, and gout) obtained the highest mean predictive improvement but also with high variance of 2.66-fold (95% CI: [1.30; 4.01]; P-value = 0.01) (Supplementary Table 4). In South Asian ancestry, the mean predictive improvement was highest in “other conditions” (including asthma, migraine, current smoker, and rheumatoid arthritis) type of 2.10-fold (95% CI: [0.787; 3.405]; P-value = 0.1). Biochemistry demonstrated the smallest improvement of 1.23-fold (95% CI: [1.15; 1.31]; P-value = 5.8 × 10^−9^). We note that PRSmix and PRSmix+ improve prediction accuracy for all traits (Supplementary Table 2 and 3). The large variance could be due to the wide range of improvement and the small number of traits in each subtype.

**Figure 4.**
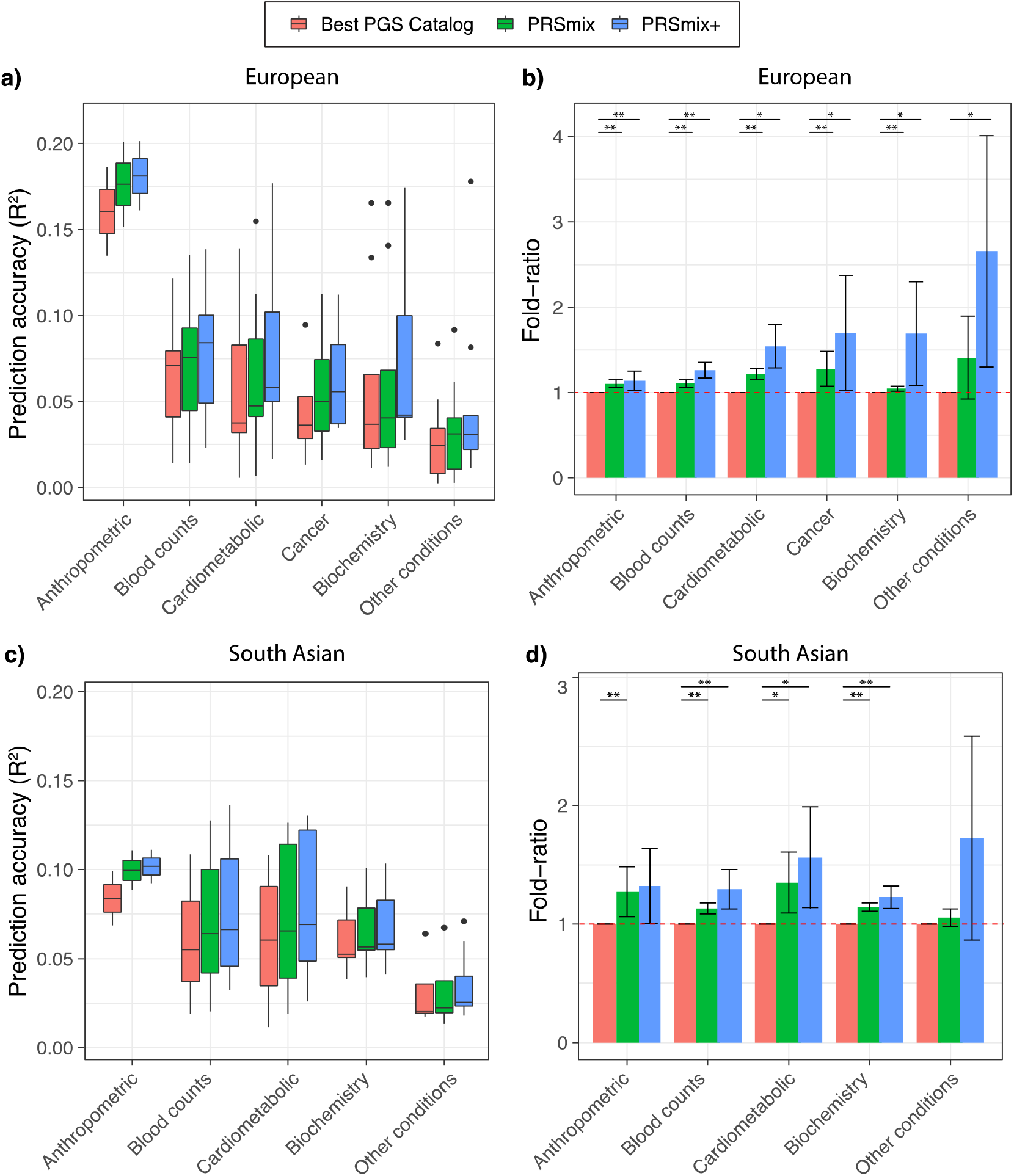
Prediction accuracy and improvement across various types of traits in the European and South Asian ancestry. We classified the traits into 6 main categories for European ancestry in the *All of Us* cohort and 5 categories for South Asian ancestry in the Genes & Health cohort due to the low prevalence of cancer traits in Genes & Health. The prediction accuracies, **(a)** and **(c)**, are estimated as partial R^2^ and liability R^2^ for continuous traits and binary traits, respectively. The relative improvements, **(b)** and **(d)**, are estimated as the fold-ratio between the prediction accuracies of PRSmix and PRSmix+ against the best PGS Catalog. The order on the axis followed the decrease in the prediction accuracy of PRSmix+. The boxplots in **(a)** and **(c)** show the first to the third quartile of prediction accuracies for 47 traits and 32 traits in European and South Asian ancestries, respectively. The whiskers reflect the maximum and minimum values within the 1.5 × interquartile range for each group. The bars in **(b)** and **(d)** represent the mean prediction accuracy across the traits in that group and the whiskers demonstrate 95% confidence intervals. The red dashed line in (b) and (d) represents the ratio equal to 1 as a reference for comparison with the best PGS Catalog score. The asterisk (*) and (**) indicate P-value < 0.05 and P-value < 0.05 / number of traits in each type with a two-tailed paired t-test, respectively.

### Comparison with previous combination methods

There have been several studies proposed to incorporate multiple traits to improve prediction accuracy of the target trait^8,19,20^. For example, wMT-SBLUP^19^ created a weighted index for correlated PRSs and required the input sample sizes, genetic correlation and heritability across all pairs of traits from GWAS summary statistics to be determined. Krapohl et al.^20^ and Albinana et al.^13^ combined PRSs using scores estimated from LDpred2^5^. Here we benchmarked PRSmix and PRSmix+ against the previous methods using summary statistics with a pre-defined set of correlated traits to the main outcomes and an extension of scores generated by different methods from PGS Catalog (Fig. 5).

**Figure 5.**
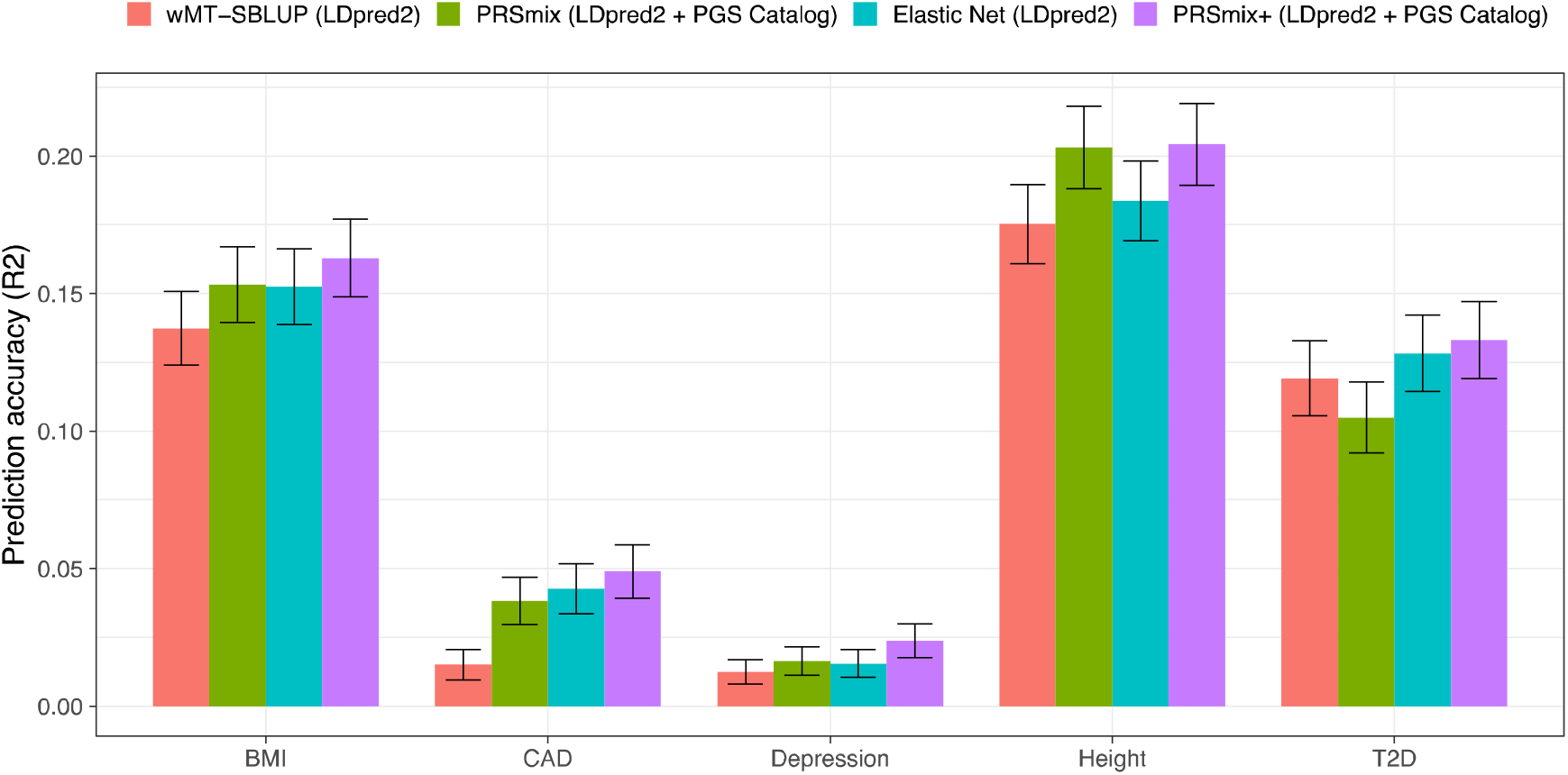
Benchmarking previous methods with PRSmix and PRSmix+. LDpred2-auto was used as the baseline method to input in the methods. 5 traits from Maier et al.^19^ and 26 publicly available GWAS for European ancestry were curated. The components of each combination method are denoted in parentheses. wMT-SBLUP was conducted with the input of sample sizes from the GWAS summary statistics and heritabilities and genetic correlation between all pairs of traits using LD score regression. PRSmix (LDpred2 + PGS Catalog) combined target trait-specific scores within 26 scores and PGS Catalog. Elastic Net (LDpred2) was performed using Elastic Net with all scores from 26 traits generated with LDpred2-auto. PRSmix+ (LDpred2 + PGS Catalog) was conducted using 26 scores from LDpred2-auto and scores from all traits obtained from PGS Catalog. Partial R2 and liability R2 were used for continuous traits and binary traits, respectively. The whiskers demonstrate 95% confidence intervals of mean prediction accuracy. BMI, Body mass index; CAD, coronary artery disease; T2D, type 2 diabetes. GWAS, genome-wide association study.

We first observed that integrating scores by Elastic Net with scores from pre-defined traits improved prediction accuracy compared to wMT-SBLUP ranging between 1.08-fold (95% CI: [1.03; 1.12]; P-value after FDR correction = 0.36) for T2D and 2.87-fold (95% CI: [1.58; 4.15]; P-value = 0.006) for CAD (Supplementary Table 5 and Supplementary Table 6). PRSmix+, with scores from both pre-defined traits and PGS Catalog, demonstrated a consistent boost in prediction accuracy compared to wMT-SBLUP between 1.12-fold (95% CI: [1.02; 1.21]; P-value = 0.016) for T2D and 3.27-fold (95% CI: [2.1; 4.44]; P-value = 2.6 × 10^−4^) for CAD. PRSmix+ equipped with both LDpred2-auto and PGS Catalog scores also outperformed the Elastic Net combination of LDpred2 scores best observed with 1.6-fold (95% CI: [1.31; 1.89]; P-value = 1.1 × 10^−4^) for depression. Interestingly, height, a highly polygenic trait^21^, demonstrated has the similarly best performance under a trait-specific combination (PRSmix with trait-specific LDpred2-auto and PGS Catalog scores) and PRSmix+ equipped with both LDpred2-auto and PGS Catalog scores (Fig. 5). Employing pleiotropic effects only provided a small improvement with height (Supplementary Table 6). On the other hand, T2D demonstrated that all methods of cross-trait combinations provided a significant improvement over the trait-specific combination (Fig. 5).

### Clinical utility for coronary artery disease

To evaluate the utility of the proposed methods, we assessed the PRSmix and PRSmix+ for coronary artery disease (CAD), which is the leading cause of disability and premature death among adults^22–24^. The single best CAD PRSs (PRS_CAD_) s from the PGS Catalog in the training set were from Koyama S. et al^25^. and Tamlander M. et al.^26^ in European and South Asian ancestries, respectively (Supplementary Fig. 5 and Supplementary Fig. 6). In the testing set, liability R^2^ with Koyama S et al. for European ancestry was 0.03 (95% CI: [0.03; 0.04]; P-value < 2 × 10^−16^) and with Tamlander M. et al. for South Asian ancestry was 0.006 (95% CI: [0.003; 0.009]; P-value = 2.39 × 10^−4^) (Fig. 6).

**Figure 6.**
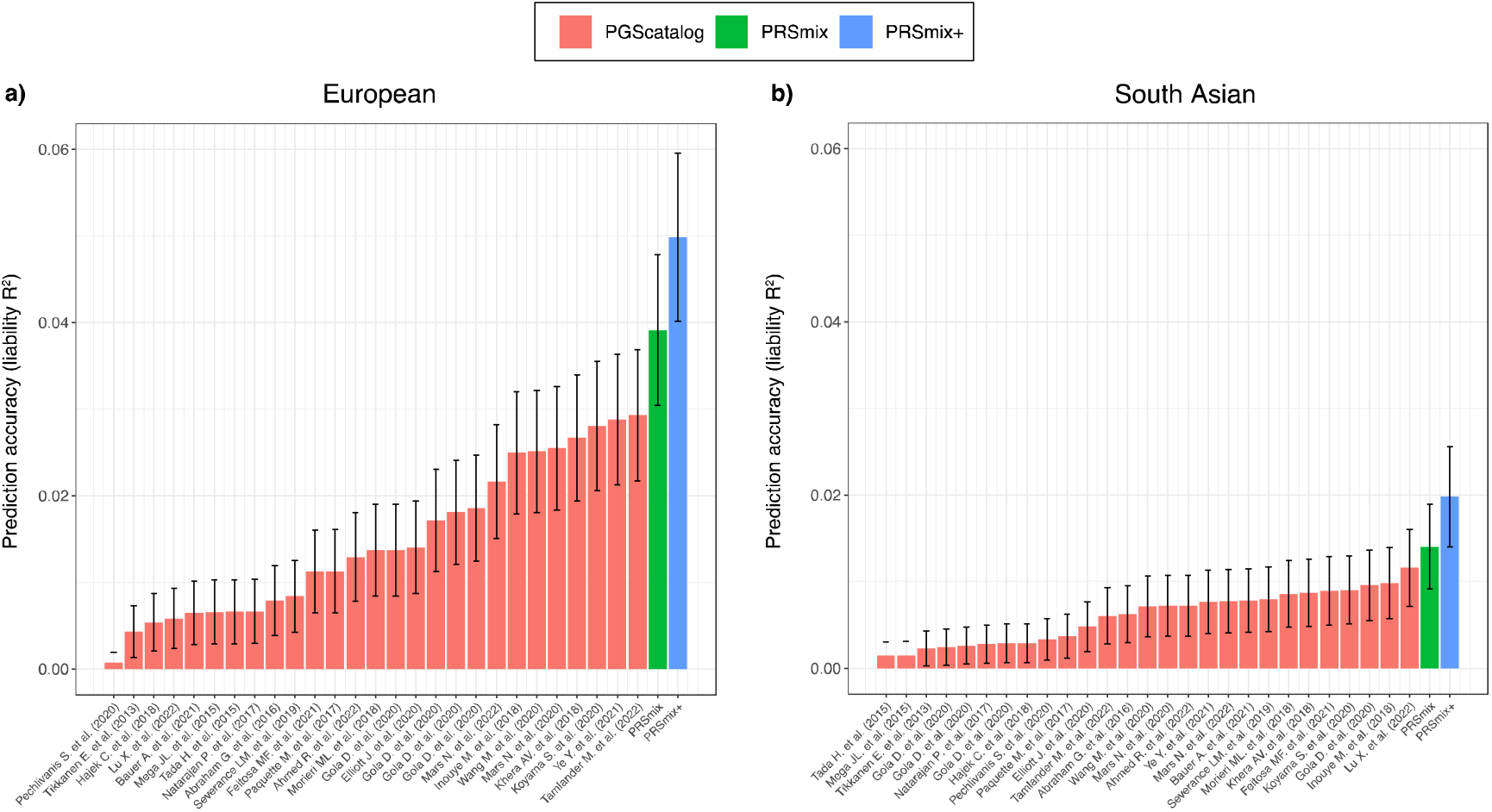
Comparison of prediction accuracies with PRSmix, PRSmix+ and CAD PRS from PGS Catalog. PRSmix was computed as a linear combination of CAD PRS and PRSmix+ was computed as a linear combination of all significant PRS obtained from the PGS Catalog. The PRSs were evaluated in the testing set with liability R^2^ in the (**a)** European ancestry from the All of Us cohort and **b)** South Asian ancestry from the Genes & Health cohort. The bars indicate the mean prediction accuracy and the whiskers show 95% confidence intervals. CAD, coronary artery disease.

Subsequently, we assessed the clinical utility of the integrative model with PRS and established clinical risk factors, including age, sex, total cholesterol, HDL-C, systolic blood pressure, BMI, type 2 diabetes, current smoking status versus the traditional model with clinical risk factors. (Fig. 7 and Supplementary Table 7). In European ancestry, the CAD PRSmix+ integrative score improved the continuous net reclassification of 35% (95% CI: [26%; 45%]; P-value < 2 × 10^−16^) compared to PRSmix (30%; 95% CI: [21%; 38%]; P-value = P-value < 2 × 10^−16^) and the best PRS from the PGS Catalog (28%; 95% CI: [19%; 38%]; P-value < 2 × 10^−16^). In South Asian ancestry, the integrated score with PRSmix+ showed significant continuous net reclassification of 27% (95% CI: [16%; 38%]; P-value = 6.07 × 10^−7^) compared to PRSmix (15%; 95% CI: [9%; 20%]; P-value = 7.18 × 10^−6^) and the best PGS Catalog (7%; 95% CI: [1%; 13%]; P-value = 0.02). Our results also demonstrated an improvement in net reclassification for models without clinical risk factors (Supplementary Table 7).

**Figure 7.**
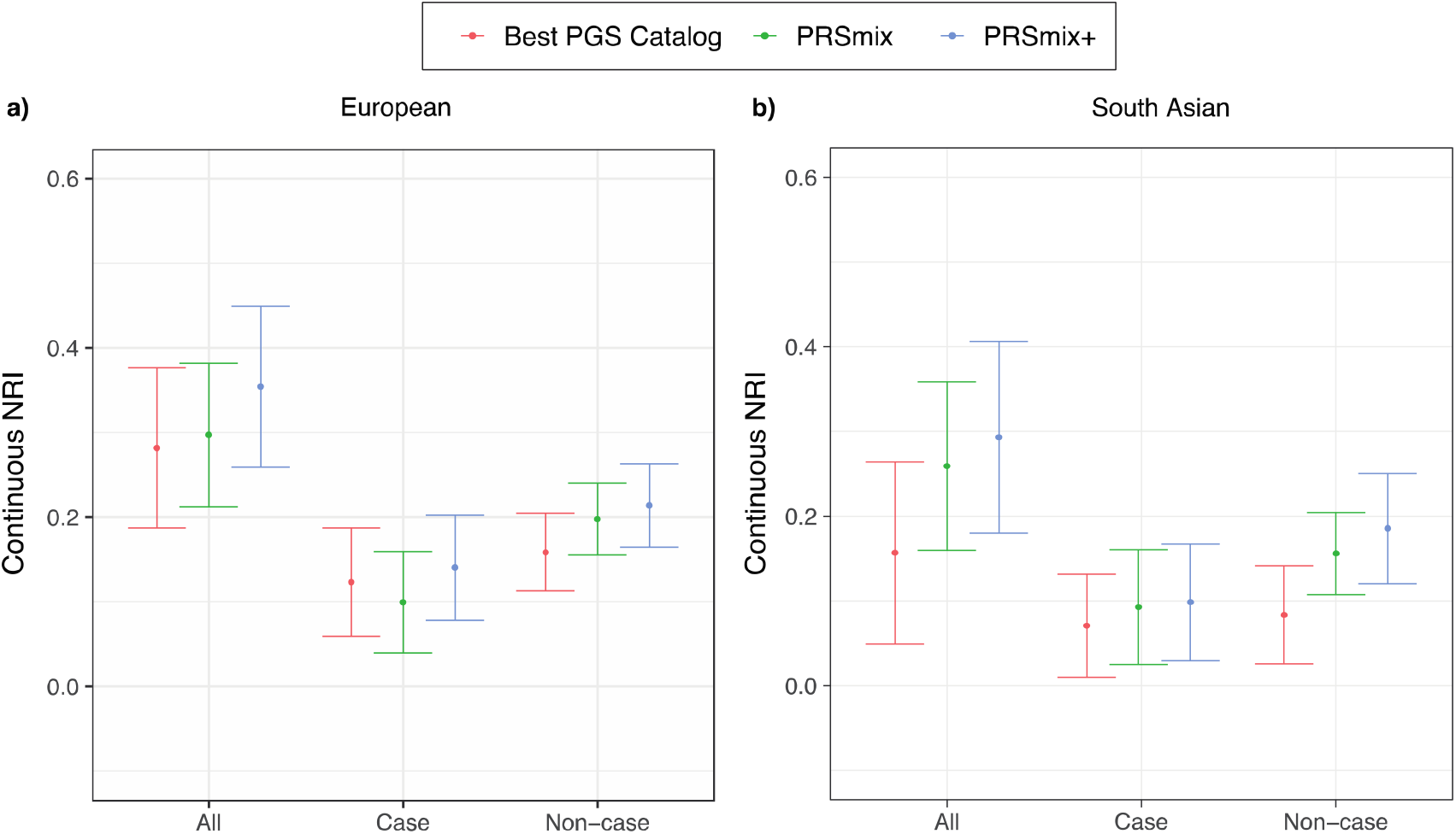
Net reclassification improvement (NRI) for coronary artery disease with the addition of polygenic risk scores to the baseline model in European and South Asian ancestries. The baseline model for risk prediction includes age, sex, total cholesterol, HDL-C, systolic blood pressure, BMI, type 2 diabetes, and current smoking status. We compared the integrative models with PGS Catalog, PRSmix, and PRSmix+ in addition to clinical risk factors versus the baseline model with only factors. The points indicate the mean estimate for continuous NRI and the whiskers indicate 95% confidence intervals estimated from 500 bootstraps. HDL-C: High-density lipoprotein; BMI: Body mass index. NRI: Net Reclassification Improvement.

We assessed the incremental area under the curve (AUC) between the full model of PRS and covariates and the null model with only covariates (Supplementary Table 8). PRSmix+ demonstrated an incremental AUC of 0.02 (95% CI: [0.018; 0.02]; P-value < 2.2×10^−16^) and 0.008 (95% CI: [0.007; 0.009]; P-value<2.2×10^−16^) in European and South Asian ancestries, respectively. PRSmix obtained an incremental AUC of 0.016 (95% CI: [0.016; 0.017]; P-value < 2.2×10^−16^) and 0.006 (95% CI: [0.005; 0.007]; P-value < 2.2×10^−16^) in European and South Asian ancestries, respectively. The best PGS Catalog had the smallest incremental AUC of 0.012 (95% CI: [0.011; 0.013]; P-value<2.2×10^−16^) and 0.003 (95% CI: [0.002; 0.003]; P-value < 2.2×10^−16^) in European and South Asian ancestries, respectively.

We also compared the risks for individuals in the top decile versus the remaining population (Supplementary Table 9). For European ancestry, an increased risk with OR per 1-SD of the best PGS Catalog, PRSmix and PRSmix+ were 1.43 (95% CI: [1.30-1.57]; P-value < 2.2×10^−16^), 1.60 (95% CI: [1.45-1.76]; P-value < 2.2×10^−16^) and 1.74 (95% CI = [1.58; 1.91]; P-value < 2.2×10^−16^), respectively. The top decile of PRSmix+ compared to the remaining population demonstrated an increased risk of OR = 2.53 (95% CI: [1.96; 3.25]; P-value =8.64 × 10^−13^). The top decile for the best PGS Catalog versus the remainder was OR = 1.67 (95% CI: [1.27; 2.19]; P-value = 2 × 10^−4^). For South Asian ancestry, an increased risk with OR per 1-SD of the best PGS Catalog, PRSmix and PRSmix+ was 1.24 (95% CI: [1.13; 1.37]; P-value < 1.52×10^−16^), 1.39 (95% CI: [1.33; 1.46]; P-value < 2.2 × 10^−16^), 1.40 (95% CI: [1.27; 1.55]; P-value < 2.2×10^−16^) and 1.50 (95% CI = [1.36; 1.66]; P-value < 2.2×10^−16^), respectively. In South Asian ancestry, PRSmix+ demonstrated an OR of 2.34 (95% CI: [1.79; 3.05]; P-value = 4.22 × 10^−10^), and with the best PGS Catalog, OR was 1.73 (95% CI: [1.30; 2.28]; P-value = 1.31 × 10^−4^) for the top decile versus the remaining population.

Moreover, we observed that there is a plateau of improvement for PRSmix from the training size of 5000 in both European and South Asian ancestries (Supplementary Fig. 7), which aligned with our simulations (Fig. 2a and 2b). Our results demonstrated the generalization of our combination methods across diverse ancestries to improve prediction accuracy. With PRSmix+, our empirical result showed that there was a modest improvement with training sample sizes larger than 5,000.

Finally, we conducted phenome-wide association studies (PheWAS) in *All of Us* between PRS_CAD_ with 1815 phecodes to compare the pleiotropy of PRS and assess the relationship between CAD PRS and disease phenotypes given the inherent use of pleiotropy in development (Supplementary Table 10). As expected, PRSmix+ had a stronger association for coronary atherosclerosis relative to the single best PRS from the PGS Catalog. PRSmix+ associations with cardiometabolic risk factors were significantly greater with averaged fold-ratio = 1.10 (95% CI: [1.09-1.12]; P-value with paired T test =1.07 × 10^−25^) and 1.07 (95% CI: [1.05-1.081]; P-value = 4.8 × 10^−13^) for circulatory system and endocrine/metabolic system (Supplementary Table 11). The PheWAS result for PRSmix+ aligned with the list of traits from the selected PRS (Supplementary Table 10).

## DISCUSSION

In this paper, we propose a trait-specific framework (PRSmix), and cross-trait framework (PRSmix+) to leverage the combined power of existing scores. We performed and evaluated our method using the *All of Us* and Genes & Health cohorts showcasing a framework to develop the most optimal PRS for a given trait in a target population leveraging all existing PRS. Across 47 traits in All of Us cohort and 32 traits in the Genes & Health cohort with either continuous traits or binary traits with prevalence > 2%, we demonstrated substantial improvement in average prediction R^2^ by using a linear combination with Elastic Net. The empiric observations are concordant with simulations. To our knowledge, there has been a number of emerging studies to combine PRS, but there is a limited number of frameworks that comprehensively evaluate, harmonize, and leverage the combination of these scores^8,13,27^. Our studies permit several conclusions for the development, implementation, and transferability of PRS.

First, externally derived and validated PRS are generally not the most optimal PRS for a given cohort. Consistent with other risk predictors, recalibration within the ultimate target population improves performance^28^. By leveraging the PGS Catalog, our work carefully harmonizes the risk alleles to estimate PRS across all scores and provides newly estimated per-allele SNP effects (provided to the PGS Catalog) to assist the interpretability of the models.

Second, previous studies selected an arbitrary training sample size to estimate the mixing weights, which may lead to a poor power of the combination frameworks and inaccurate estimate of sampling variance^10^. We assessed the expected sample sizes to estimate the mixing weights via simulations and real data. Our results demonstrated that while low heritability traits benefit the most, they require a greater training sample size.

Third, we leveraged all PRS, including those not trained on the primary trait, to systematically optimize PRS for a target cohort. We showed that PRSmix improved the prediction by combining the scores matching the outcome trait. In addition, we showed that PRSmix+ was able to leverage the power of cross-traits, which highlighted the contribution of pleiotropic effects to enhance PRS performance. We leverage prior work demonstrating the effects of pleiotropy on complex traits^15,29,30^.

Fourth, we demonstrated that our method outperformed previous methods combining scores. We showed that PRSmix+ outperformed wMT-SBLUP^19^ using a limited number of correlated traits. wMT-SBLUP required GWAS’s sample sizes, heritability, and genetic correlation between all traits. LDpred2-auto required GWAS summary statistics and initialized heritability and proportion of causal SNPs. Krapohl et al.^20^ and Abraham et al.^8^ proposed to use Elastic Net to combine the scores developed from summary statistics, and correlated traits were selected with prior knowledge. However, these strategies consider scores developed from a particular methods using predefined summary statistics. Our framework utilizes all PRSs available in the PGS Catalog which were optimized for their target traits. Additional summary statistics and PRS scores could be added to further enhance the models. We let our Elastic Net model penalize the component PRSs without the need for prior knowledge. Elastic Net can select PRSs to include and efficiently handle multi-collinearity^31–33^. Furthermore, PRSmix and PRSmix+ only required a set of SNPs effect to estimate the PRSs and estimated the prediction accuracy to the target trait to select the best scores for the combination. Additionally, compared to the preselected traits for stroke by Abraham et al.^8^ we also observed that our method could identify more related risk factors to include compared to previous work conducted on stroke (Supplementary Fig. 8). Therefore, our method is more comprehensive in an unbiased way in terms of choosing the risk factors and traits to include with empirically improved performance.

Fifth, greater performance is observed even for non-European ancestry groups underrepresented in GWAS and PRS studies. We empirically demonstrate the value of training and incorporating pleiotropy with all available PRS to improve performance, including multiple metrics of clinical utility for CAD prediction in multiple ancestries. In South Asian ancestry, we observed that PRSmix and PRSmix+ demonstrated a significant improvement with the best improvement for CAD. Of note for CAD, the relative improvements in South Asian ancestry were higher than in European ancestry for PRSmix and equivalent for PRSmix+. Transferability of PRS has been shown to improve the clinical utility of PRS in non-European ancestry^16,34^. Although the prediction accuracy for South Asian ancestry is still limited, our results highlighted the transferability of predictive improvement with PRSmix and PRSmix+ to South Asian ancestry. We anticipate that ongoing and future efforts to improve our understanding of the genetic architecture in non-European ancestries will further improve the transferability of PRS across ancestry.

Lastly, traits with low heritability or generally low-performing single PRS benefit the most from this approach, especially with PRSmix+, such as migraine in both European and South Asian ancestries. Additionally, our results showed that pleiotropic effects play an important role in understanding and improving prediction accuracies of complex traits. However, anthropometric traits, which are highly polygenic^35^ and have good predictive performance using the best PGS Catalog, also showed improvement with the combination framework in both European and South Asian ancestries.

Given that PRSmix+ outperformed PRSmix, one might consider if there is a reason to use PRSmix instead of PRSmix+. We observed that in cases of highly heritable traits or high performance with a single PRS, there was only marginal improvement of PRSmix+ over PRSmix. In this scenario, PRSmix could provide similar predictive performance while being less time-consuming because trait-specific PRS inputs only are required. However, for traits with lower heritability PRSmix+ shows a marked improvement over PRSmix and would be preferred. Wang et al.^36^ showed that the theoretical prediction accuracy of the target trait using the PRS from the correlated trait is a function of genetic correlation, heritability, number of genetic variants and sample size. Future directions could include defining the minimum parameters required for the performance of the PRSmix+ model to improve on single trait-specific PRS.

Our work has several limitations. First, the majority of scores from PGS Catalog were developed in European ancestry populations. Further non-European SNP effects will likely improve the single PRS power, which may in turn, also improve the prediction accuracy of our proposed methods. Second, the Elastic Net makes a strong assumption that the outcome trait depends on a linear association with the PRS and covariates. However, a recent study demonstrated there is no statistical significance difference between linear and non-linear combinations for neuropsychiatric disease^13^. Third, we did not validate the mixing weights in an independent cohort. We expect that in the future, there will be emerging large independent biobanks, but prior non-genetic work demonstrates the value of internal calibration for optimal risk prediction. Fourth, we estimated the mixing weights for each single SNP as a mixing weight of the PRS. Future studies could consider linkage disequilibrium between the SNPs and functional annotations of each SNP. Fifth, our frameworks were conducted on binary traits with a prevalence > 2%. Additional combination PRS models are emerging that seek to use preexisting genotypic data from genetically related, but low prevalence conditions, to improve the prediction accuracy of rare conditions^13^. Sixth, the baseline demographic characteristics (i.e., age, sex, social economic status) in the target cohort might limit the validation and transferability of PRS^37^. Although these factors were considered by using a subset of the target cohorts as training data, it is necessary to have PRS developed on similar baseline characteristics. Lastly, with the expanding of all biobanks, there might be no perfect distinction between the samples deriving PRS and the testing cohort, future studies may consider the potential intersection samples to train the linear combination.

In conclusion, our framework demonstrates that leveraging different PRS either trait-specific or cross-trait can substantially improve model stability and prediction accuracy beyond all existing PRS for a target population. Importantly, we provide software to achieve this goal in independent cohorts.

## METHODS

### Data

#### The All of Us Research Program

The *All of Us* Research Program is a longitudinal cohort continuously enrolling (starting May 2017) U.S. adults ages 18 years and older from across the United States, with an emphasis on promoting inclusion of diverse populations traditionally underrepresented in biomedical research, including gender and sexual minorities, racial and ethnic minorities, and participants with low levels of income and educational attainment.^38^ Participants in the program can opt-in to providing self-reported data, linking electronic health record data, and providing physical measurement and biospecimen data.^39^ Details about the *All of Us* study goals and protocols, including survey instrument development,^40^ participant recruitment, data collection, and data linkage and curation were previously described in detail.^39,41^

Data can be accessed through the secure *All of Us* Researcher Workbench platform, which is a cloud-based analytic platform that was built on the Terra platform.^42^ Researchers gain access to the platform after they complete a 3-step process including registration, completion of ethics training, and attesting to a data use agreement attestation.^43^ *All of Us* uses a tiered approach based on what genomic data is accessible through the Controlled Tier, and includes both whole genome sequencing (WGS), genotyping array variant data in multiple formats, as well as variant annotations, access to computed ancestry, and quality reports.^44^ This study includes data on the 98,600 participants with (WGS) data in the *All of Us* v6 Curated Data Repository release. Participant data in this data release was collected between May 6, 2018 and April 1, 2021. This project is registered in the *All of Us* program under the workspace name “Polygenic risk score across diverse ancestries and biobanks.”

#### The Genes & Health Biobank

Genes & Health is a community-based genetics study enrolling British South Asian, with an emphasis on British Bangladeshi (two-thirds) and British Pakistani (remaining) people, with a goal of recruiting at least 100,000 participants. Currently, over 52,000 participants have enrolled since 2015. All participants have consented for lifelong electronic health record access and genetic analysis. The study was approved by the London South East National Research Ethics Service Committee of the Health Research Authority. 97.4% of participants in Genes & Health are in the lowest two quintiles of the Index of Multiple Deprivation in the United Kingdom. The cohort is broadly representative of the background population with regard to age, but slightly over-sampled with females and those with medical problems since two-thirds of people were recruited in healthcare settings such as General Practitioner surgeries^45^.

#### The Polygenic Score (PGS) Catalog

Polygenic risk scores were obtained from the Polygenic Score (PGS) Catalog^17^, which is a publicly accessible resource cataloging published PRS, including the metadata. The metadata provides information describing the computational algorithms used to generate the score, and performance metrics to evaluate a PRS^17^. At the time of this study, over 2,600 PRS were cataloged in the PGS Catalog (version July 18, 2022) designed to predict 538 distinct traits.

### Clinical Outcomes

Clinical phenotypes were curated using a combination of electronic health record data, direct physical measurements, and/or self-reported personal medical history data, from the *All of Us* v6 Data Release as detailed in Supplementary Table 16. Individuals in the Genes and Health cohort were also curated with similar definitions based on ICD10, SNOMED and operation codes (Supplementary Table 17). Traits with the best performing single trait-specific PRS with power < 0.95 such as hemoglobin, sleep apnea, and depression were removed. Binary traits with a prevalence < 2% were removed.

### A linear combination of scores

We proposed PRSmix to combine PRS of outcome traits and PRSmix+ to combine high-power PRS (defined in the following subsection) from all traits obtained from PGS Catalog. The linear combination was conducted by using an Elastic Net algorithm from the “*glmnet*” R package^46^ (version 4.1) to combine the estimated PRS. First, we randomly split the independent cohorts into 80% of training and 20% testing. The PRS in the training set was standardized with mean 0 and variance 1. Before conducting linear combination, we first evaluated the performance of each individual PRS by their power and P-value (see below). An Elastic Net algorithm was used with 5-fold cross-validation and default parameters to estimate the mixing weights of each PRS. The mixing weights were then divided by the corresponding original standard deviation of the PRS in the training set.

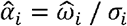

Where 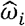 and *σ*_i_ is the mixing weight estimated from the Elastic Net and standard deviation of PRS_i_ in the training set, respectively. 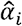 is the adjusted mixing weight for PRS_i_. To derive the per-allele effect sizes from the combination framework, we multiplied the SNP effects with the corresponding adjusted mixing weights:

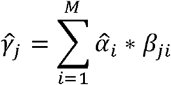

Where 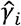 is the adjusted effect size of SNP_j_ and *β*_*ij*_ is the original effect sizes of SNP_j_ in PRS_i_. We set *β*_*ij*_ = 0 if SNP_j_ is not in PRS_i_. The adjusted effect sizes were then utilized to calculate the final PRS.

The mixing weights for PGS Catalog scores for PRSmix and PRSmix+ in European ancestry are provided in Supplementary Table 12 and Supplementary Table 13, respectively. For South Asian ancestry, the mixing weights for PRSmix and PRSmix+ in European ancestry are provided in Supplementary Table 14 and Supplementary Table 15, respectively.

### Power and variance of PRS accuracy

We selected high-power PRS to conduct the combination by assessing the power and variance of prediction accuracy. The power of PRS can be estimated based on the power of the two-tailed test of association as follow^3,47^:

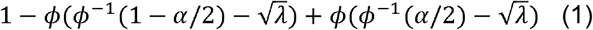

where *ϕ* is the Chi-squared distribution function, α is the significance level, and λ is the non-centrality parameter which can be estimated as

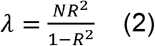

where N, *R*^2^ is the sample size and estimated prediction accuracy in the testing set, respectively. *R*^2^ can be estimated as partial *R*^2^ or liability *R*^2^ for continuous traits and binary traits, respectively. Briefly, partial *R*^2^ compared the difference in goodness-of-fit between a full model with PRS and covariates including age, sex, and first 10 PCs, and a null model with only covariates. Additionally, for binary traits, liability *R*^2^ was estimated with the disease prevalence approximated as the prevalence in the samples. The theoretical variance and standard error of *R*^2^ can be estimated as follow^48–50^:

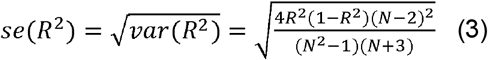

Therefore, we can analytically estimate the confidence interval of prediction accuracy for each of the score. We selected high-power scores defined as power > 0.95 with P-value <= 0.05 or P-value <= 1.9 × 10^−5^ (0.05/2600) for the combination with Elastic Net.

To compare the improvement, for instance between PRSmix and the best PGS Catalog, we estimate the mean fold-ratio of R^2^ across different traits with its 95% confidence interval and evaluated the significance difference from 1 using a two-tailed paired t-test.

### Simulations

We used UK Biobank European ancestry to conduct simulations for trait-specific and cross-trait combinations. Overall, we simulated 7 traits with heritability *h*^2^ equal to 0.05, 0.1, 0.2, and 0.5. We randomly selected M=1000 causal SNPs among 1.1 million HapMap3 variants with INFO > 0.6, MAF > 0.01 and P-value Hardy-Weinberg equilibrium > 10^−7^. We removed individuals with PC1 and PC2 > 3 standard deviation from the mean. We randomly remove one in a pair of related individuals with closer than 2nd degree. The genetic components were simulated as PRSs where PRS1, PRS2, and PRS3 are considered trait-specific scores with genetic correlations are 0.8 and 0.4 for cross-trait scores. PRS4, PRS5 and PRS6 are simulated as pleiotropic effects on the outcome traits with genetic correlation equal to 0.4. The SNP effects for PRSs are simulated by a multivariate normal distribution MVN(0, *∑*) where *∑* is the covariance matrix between PRSs. The main diagonal contains the heritability of the traits as *h*^2^ / *M* and the covariance between PRSs are simulated as *r*_*g*_ *\* h*^2^ / *M* where *r*_*g*_ is the genetic correlation between PRSs (0.8 for trait-specific scores and 0.4 for cross-trait scores). The PRSs of the outcome are estimated by the weighted combination of PRS where the weights follow U(0,1). 7 phenotypes were simulated as *y* = *g* + *e,e* ~ *N*(0,1 - *h*^2^) where g is PRS and e is the residuals.

We split the simulated cohort into 3 data sets for: 1) GWAS 2) training set: training the mixing weights with a linear combination and 3) testing set: testing the combined PRS. We incorporated PRS1, PRS2 and PRS3 to assess the trait-specific PRSmix framework. We combined all 6 single PRS to evaluate the cross-trait PRSmix+ framework. We compared the fold-ratio of the R^2^ of the combined PRS to the R^2^ of best single PRS to assess the improvement of the combination strategy. To evaluate the improvement across different heritabilities, we estimated the slope of improvement per log10(N) increase of training sample sizes on the fold-ratio of predictive improvement.

### Sample and genotyping quality control

The AoU data version 5 contains more than 700 million variants from whole genome sequencing^39^. We curated European ancestry by predicted genetic ancestry with a probability > 90% provided by AoU yielding 48,112 individuals in the AoU. For variant quality control beyond AoU central efforts, we further filtered SNPs to include MAF > 0.001 which retained 12,416,130 SNPs. We performed a similar quality control for imputed genotype data for South Asian ancestry in the Genes & Health cohort with additional criteria of INFO score > 0.6 and genotype missing rate < 5%. Individuals with a missing rate > 5% were removed. Eventually, 44,396 individuals and 8,935,207 SNPs remained in Genes & Health.

### Assessment of clinical utility

We applied PRSmix and PRSmix+ for coronary artery disease as a clinical application. The phenotypic algorithm includes at least one ICD or CPT code below: ICD9 410x, 411x, 412x; ICD10 I22x, I23x, I24.1, I25.2 CPT 92920-92979 (PCI), 33533-33536, 33517-33523, 33510-33516 (CABG) or self-reported personal history of MI or CAD. CAD in Genes and Health cohort was defined with at least one ICD10 I22x, I23x, I24.1, I25 or operation codes K401, K402, K403, K404, K411, K451, K452, K453, K454, K455, K491, K492, K499, K502, K751, K752, K753, K754, K758, K759 or SNOMED codes 1755008, 22298006, 54329005, 57054005, 65547006, 70211005, 70422006, 73795002, 233838001, 304914007, 401303003, 401314000.

The category-free NRI was used to evaluate the clinical utility. NRI was calculated by adding the PRS to the baseline logistic model including age, sex, the first 10 principal components, and clinical risk factors. The clinical risk factors include total cholesterol, HDL-C, BMI, type 2 diabetes, and current smoking status or model includes only age, sex, and 10 principal components. NRI was calculated as the sum of NRI for cases and NRI for controls:

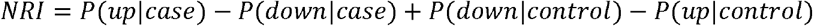

*P*(*up*|*case*) and P(*down*|*case*) estimate the proportion of cases that had higher or lower risk after classification with logistic regression, respectively. The confidence interval for NRI was estimated with 500 bootstraps. We also compared the risk increase between individuals in the top decile of PRS versus those remaining in the population. In addition to liability R^2^ to compare the PRS performance, we also used the incremental area under the curve (AUC) to compare the PRS. The incremental AUC was estimated as the difference between the AUC of models with the integrative score versus the model with only clinical variables.

### wMT-SBLUP and linear combination of LDpred2-auto derived scores

#### LDpred2-auto

LDpred2 is a Bayesian method that computes the adjusted SNP effect sizes from GWAS summary statistics. LDpred2 utilizes the SNP effect sizes as prior and incorporates LD between markers to infer the posterior effect sizes. In our work, we implemented LDpred2-auto^51^ since this method can infer heritability and the proportion of causal variants. LDpred2-auto was conducted with 800 burn-in iterations and 500 iterations. The proportion of causal variants was initialized between 10^−4^ and 0.9. Furthermore, LDpred2-auto does not require a validation set, the SNP effect sizes were averaged between scores. We used 1,138,726 HapMap3 variants that overlapped with SNPs from whole-genome sequencing data in the All of Us cohort. The LD reference panel developed from European ancestry was provided by the LDpred2-tutorial.

#### wMT-SBLUP

wMT-SBLUP^19^ calculates the mixing weights of PRS using sample sizes from GWAS summary statistics, SNP-heritability and genetic correlation. We compared wMT-SBLUP with our method using 5 traits that were originally assessed with wMT-SBLUP including CAD, T2D, depression, height, and BMI. We curated 26 publicly available GWAS summary statistics (Supplementary Table 18) and performed LDpred2-auto with quality controls suggested by Privé et al^5,51^. We used LD score regression to estimate SNP-heritability and genetic correlation across 26 traits. For each of the 5 outcome traits, we selected correlated traits with P-value of genetic correlation less than 0.05.

#### Elastic Net for linear combination

we also implemented linear combination by Elastic Net with the LDpred2-auto-derived PRSs for contributing traits since this strategy was proposed by several works^8,13,20^. We selected scores with significant variance explained (P-value<0.05) to the outcome trait and conducted Elastic Net using the *glmnet* R package^46^.

### Phenome-wide association study

We obtained the list of 1815 phecodes from the PheWAS website (last accessed December 2022)^52^. The phecodes were based on ICD-9 and ICD-10 to classify individuals. PheWAS was conducted on European ancestry only in AoU. For each phecodes as the outcome, we conducted an association analysis using logistic regression on PRS and adjusted for age, sex, and first 10 PCs. The significance threshold for PheWAS was estimated as 2.75 × 10^−5^ (0.05/1815) after Bonferroni correction.

## Supporting information

Supplementary Figure 1-8 and legends for Supplementary Table 1-18

## Data Availability

The PGS Catalog is freely available at https://www.pgscatalog.org/. Our new scores are deposited in the PGS Catalog. The All of Us and Genes & Health individual-level data is a controlled access dataset and may be granted at https://www.researchallofus.org/ and https://www.genesandhealth.org/, respectively.

## Data availability

The weights from the PRSmix and PRSmix+ scores in this manuscript have been returned to the PGS Catalog. The R package to implement PRSmix and PRSmix+ in independent datasets is at https://github.com/buutrg/PRSmix.

### Software/analyses

Analyses were performed on the AoU Researcher Workbench in Jupyter Notebook 14 using R version 4.0.0 programming language. Results are reported in compliance with the AoU Data and Statistics Dissemination Policy.

## ACKNOWLEDGEMENT

We would like to thank Alkes L. Price for critical comments for this works. L.E.H. is supported by the National Human Genome Research Institute (K08HG012221). P.N. is supported by grants from NHGRI (U01HG011719), NHLBI (R01HL142711, R01HL127564, R01HL151152), and Massachusetts General Hospital (Paul & Phyllis Fireman Endowed Chair in Vascular Medicine). The content is solely the responsibility of the authors and does not necessarily represent the official views of the National Institutes of Health.

The *All of Us* Research Program is supported by the National Institutes of Health, Office of the Director: Regional Medical Centers: 1 OT2 OD026549; 1 OT2 OD026554; 1 OT2 OD026557; 1 OT2 OD026556; 1 OT2 OD026550; 1 OT2 OD 026552; 1 OT2 OD026553; 1 OT2 OD026548; 1 OT2 OD026551; 1 OT2 OD026555; IAA #: AOD 16037; Federally Qualified Health Centers: HHSN 263201600085U; Data and Research Center: 5 U2C OD023196; Biobank: 1 U24 OD023121; The Participant Center: U24 OD023176; Participant Technology Systems Center: 1 U24 OD023163; Communications and Engagement: 3 OT2 OD023205; 3 OT2 OD023206; and Community Partners: 1 OT2 OD025277; 3 OT2 OD025315; 1 OT2 OD025337; 1 OT2 OD025276. In addition, the All of Us Research Program would not be possible without the partnership of its participants.

Genes & Health is/has recently been core-funded by Wellcome (WT102627, WT210561), the Medical Research Council (UK) (M009017), Higher Education Funding Council for England Catalyst, Barts Charity (845/1796), Health Data Research UK (for London substantive site), and research delivery support from the NHS National Institute for Health Research Clinical Research Network (North Thames). Genes & Health is/has recently been funded by Alnylam Pharmaceuticals, Genomics PLC; and a Life Sciences Industry Consortium of Bristol-Myers Squibb Company, GlaxoSmithKline Research and Development Limited, Maze Therapeutics Inc, Merck Sharp & Dohme LLC, Novo Nordisk A/S, Pfizer Inc, Takeda Development Centre Americas Inc.

We thank Social Action for Health, Centre of The Cell, members of our Community Advisory Group, and staff who have recruited and collected data from volunteers. We thank the NIHR National Biosample Centre (UK Biocentre), the Social Genetic & Developmental Psychiatry Centre (King’s College London), Wellcome Sanger Institute, and Broad Institute for sample processing, genotyping, sequencing and variant annotation. We thank: Barts Health NHS Trust, NHS Clinical Commissioning Groups (City and Hackney, Waltham Forest, Tower Hamlets, Newham, Redbridge, Havering, Barking and Dagenham), East London NHS Foundation Trust, Bradford Teaching Hospitals NHS Foundation Trust, Public Health England (especially David Wyllie), Discovery Data Service/Endeavour Health Charitable Trust (especially David Stables), NHS Digital - for GDPR-compliant data sharing backed by individual written informed consent.

Most of all we thank all of the volunteers participating in the *All of Us* Research Program and Genes & Health.

