## Supplementary Figure 1-8 and legends for Supplementary Table 1-18 for "Integrative polygenic risk score improves the prediction accuracy of complex traits and diseases"

Buu Truong et al.

###### **Supplementary Figure 1. Prediction accuracy of the best PGS catalog versus PRSmix and PRSmix+ for 47 traits in the European ancestry.**

The prediction accuracies of the best PGS Catalog are displayed on the x-axis versus PRSmix **(a)** and PRSmix+ **(b)** on the y-axis. The prediction accuracies are estimated as partial R2 and liability R2 for continuous traits and binary traits, respectively. The points represent the mean prediction and the whiskers demonstrate 95% confidence intervals. The black line represents the reference that the prediction accuracy of PGS Catalog is equal to PRSmix and PRSmix+.

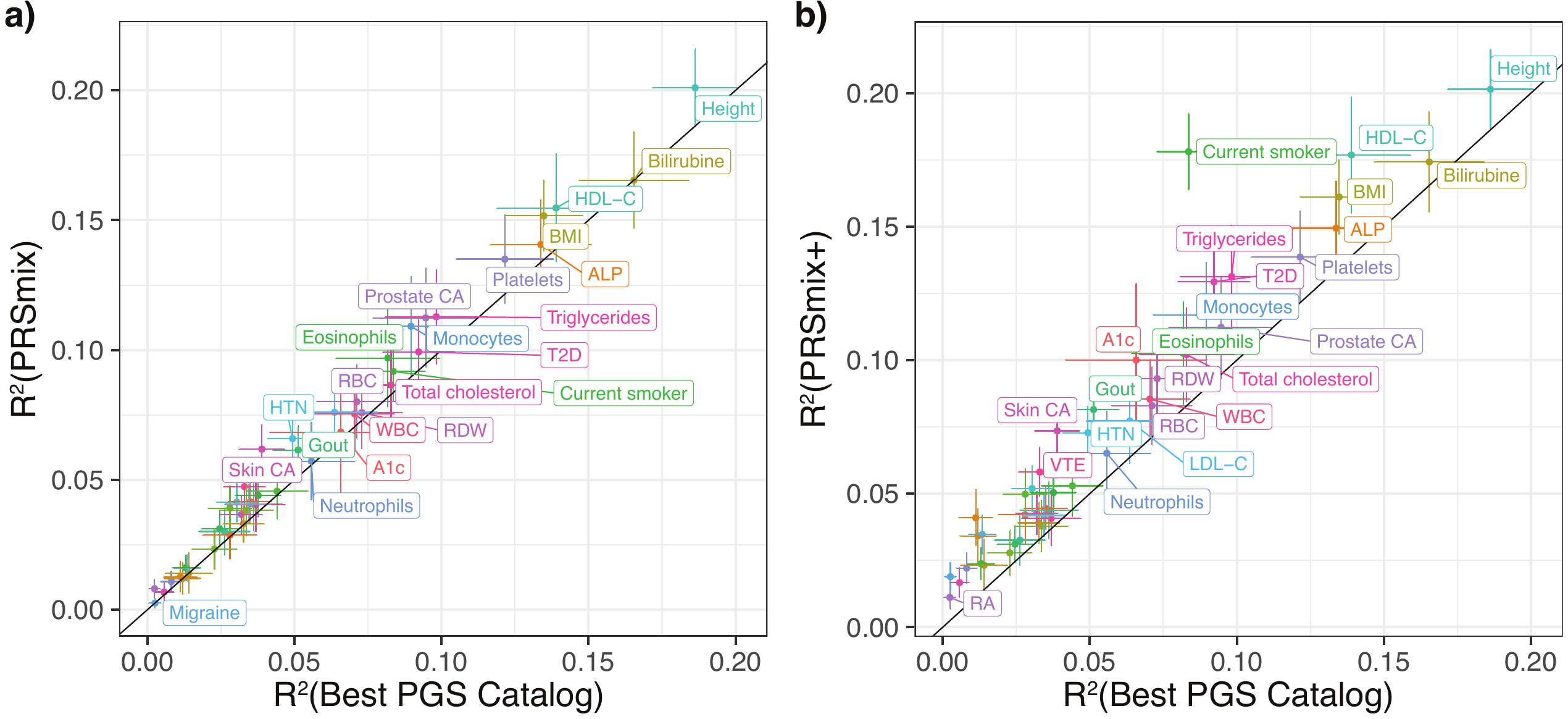

###### **Supplementary Figure 2. Prediction accuracy of the best PGS catalog versus PRSmix and PRSmix+ for 32 traits in the South Asian ancestry.**

The prediction accuracies of the best PGS Catalog are displayed on the x-axis versus PRSmix **(a)** and PRSmix+ **(b)** on the y-axis. The prediction accuracies are estimated as partial R2 and liability R2 for continuous traits and binary traits, respectively. The points represent the mean prediction and the whiskers demonstrate 95% confidence intervals. The black line represents the reference that the prediction accuracy of PGS Catalog is equal to PRSmix and PRSmix+.

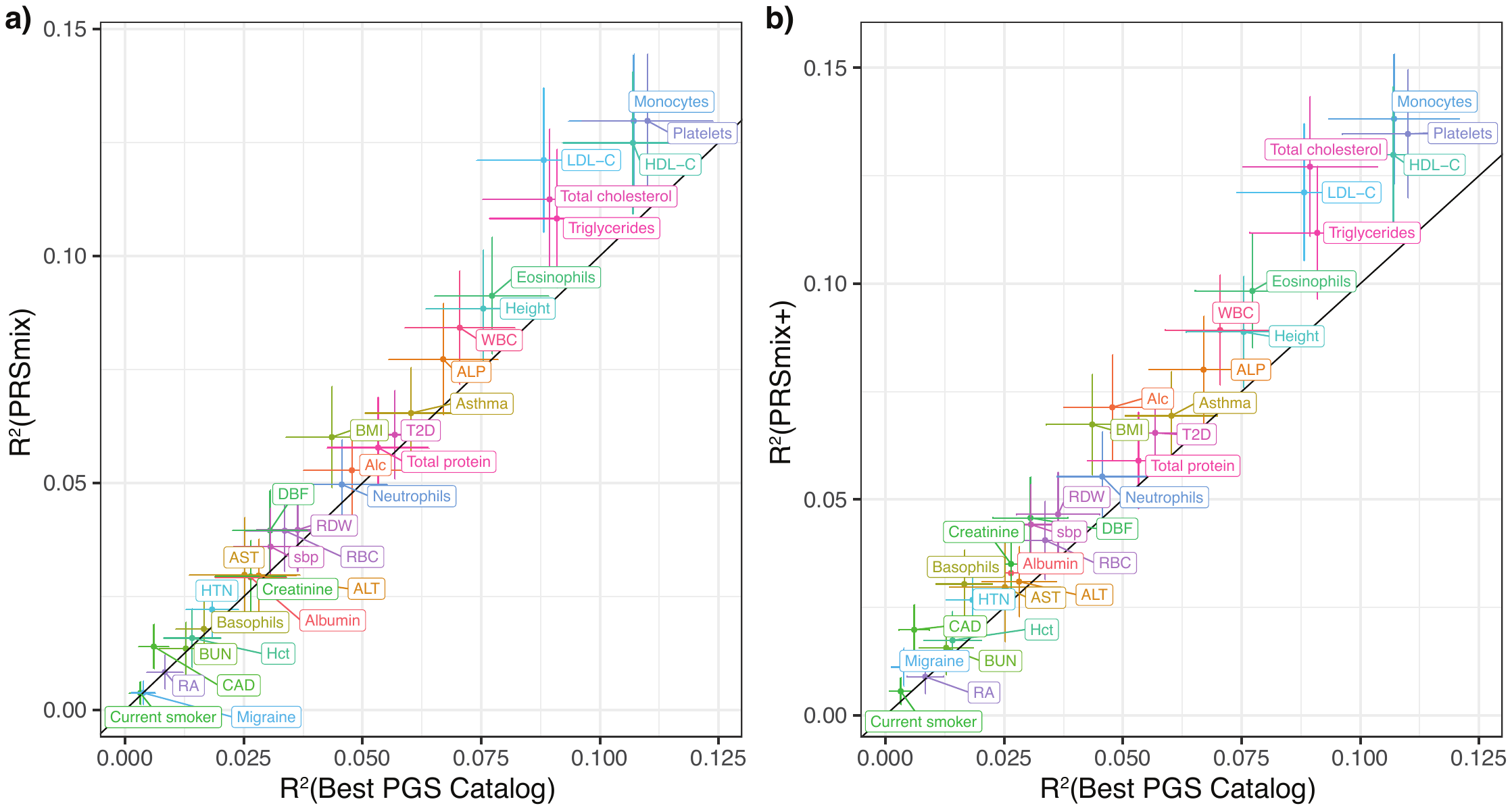

###### **Supplementary Figure 3. Prediction accuracy of the PRSmix versus PRSmix+ in the European and South Asian ancestries.**

The prediction accuracies of the PRSmix are displayed on the x-axis versus PRSmix+ on the y-axis in **a)** European ancestry and **b)** South Asian ancestry. The prediction accuracies are estimated as partial R2 and liability R2 for continuous traits and binary traits, respectively. The points represent the mean prediction and the whiskers demonstrate 95% confidence intervals. The black line represents the reference that the prediction accuracy of PGS Catalog is equal to PRSmix and PRSmix+.

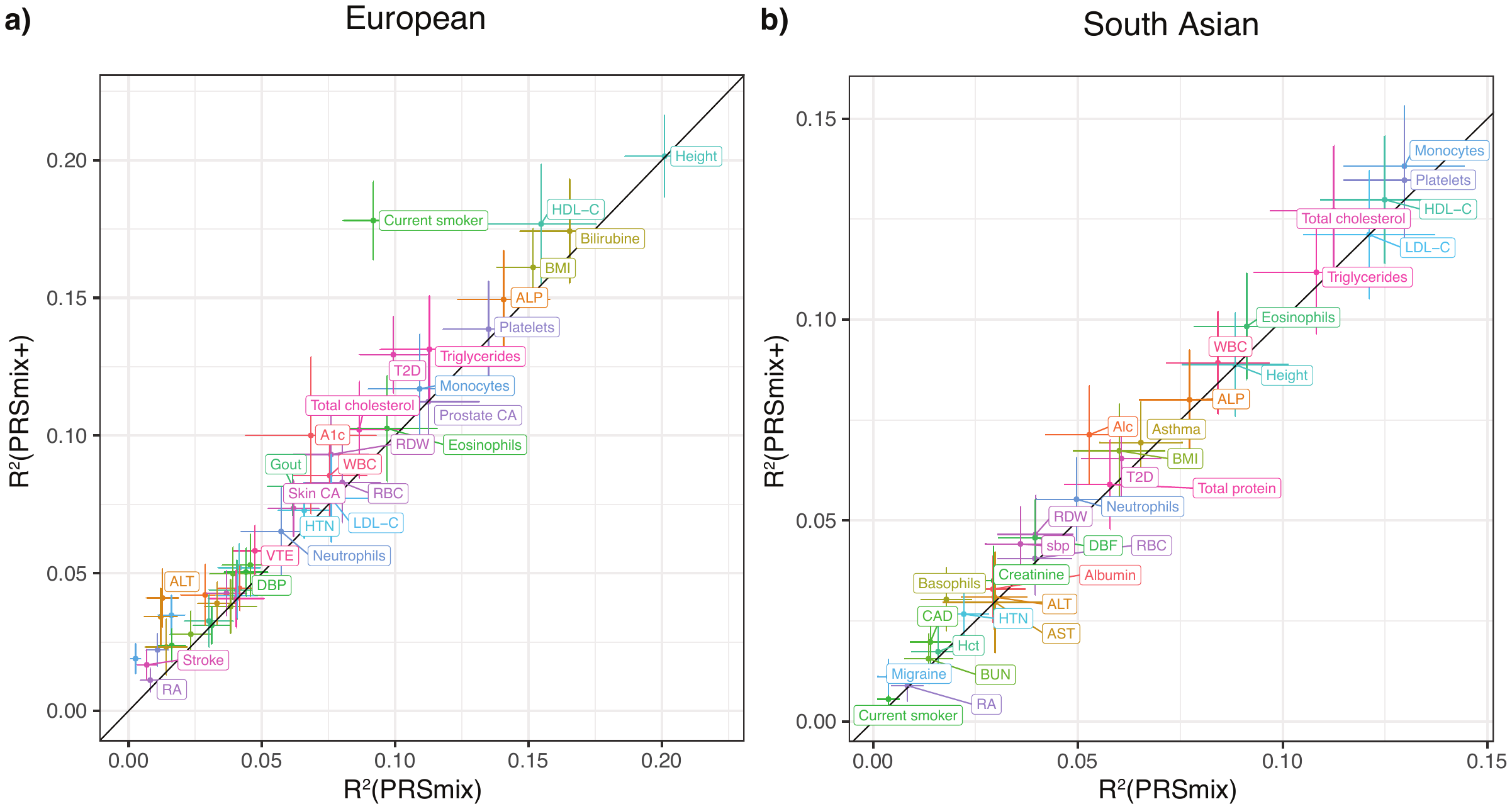

###### **Supplementary Figure 4. Predictive improvement of PRSmix and PRSmix+ in European and South Asian ancestries.**

The prediction accuracies of the best PGS Catalog are displayed on the x-axis versus ratio of R^2^_PRSmix_ and R^2^_PRSmix+_ on the y-axis. The prediction accuracies are estimated as partial R2 and liability R2 for continuous traits and binary traits, respectively. The red and blue line represents the local regression line for improvement of PRSmix and PRSmix+ over the best PGS Catalog, respectively.

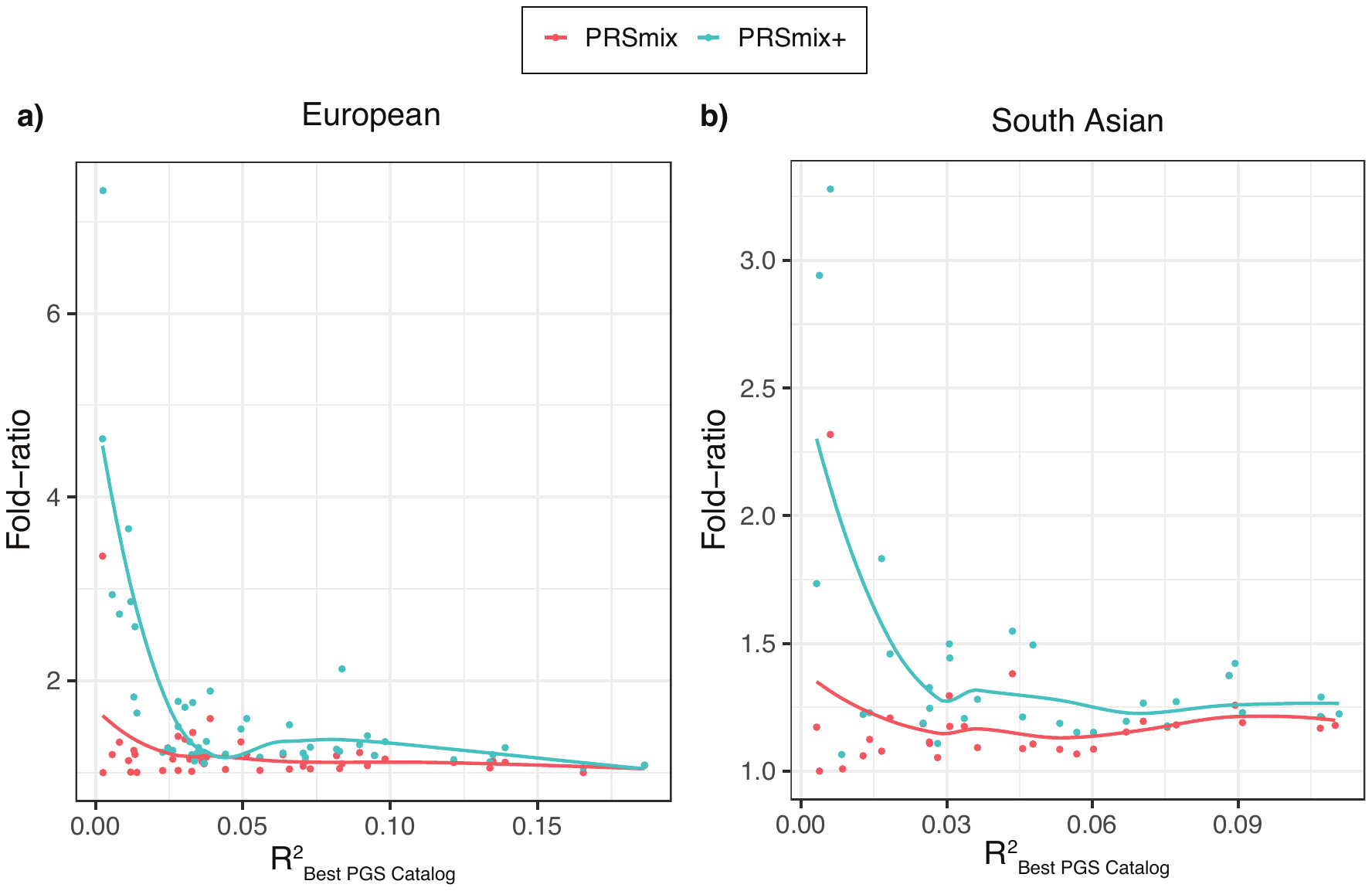

###### **Supplementary Figure 5. Prediction accuracy in the training set of the contributing PGSs to PRSmix+ of coronary artery disease in the European ancestry.**

We used Elastic Net to estimate the mixing weights of multiple PGS and report the contributing PGS with non-zero effects. Prediction accuracy was estimated with liability R2. The boxes represent the mean prediction accuracy across the traits in that group and the whiskers demonstrate 95% confidence intervals.

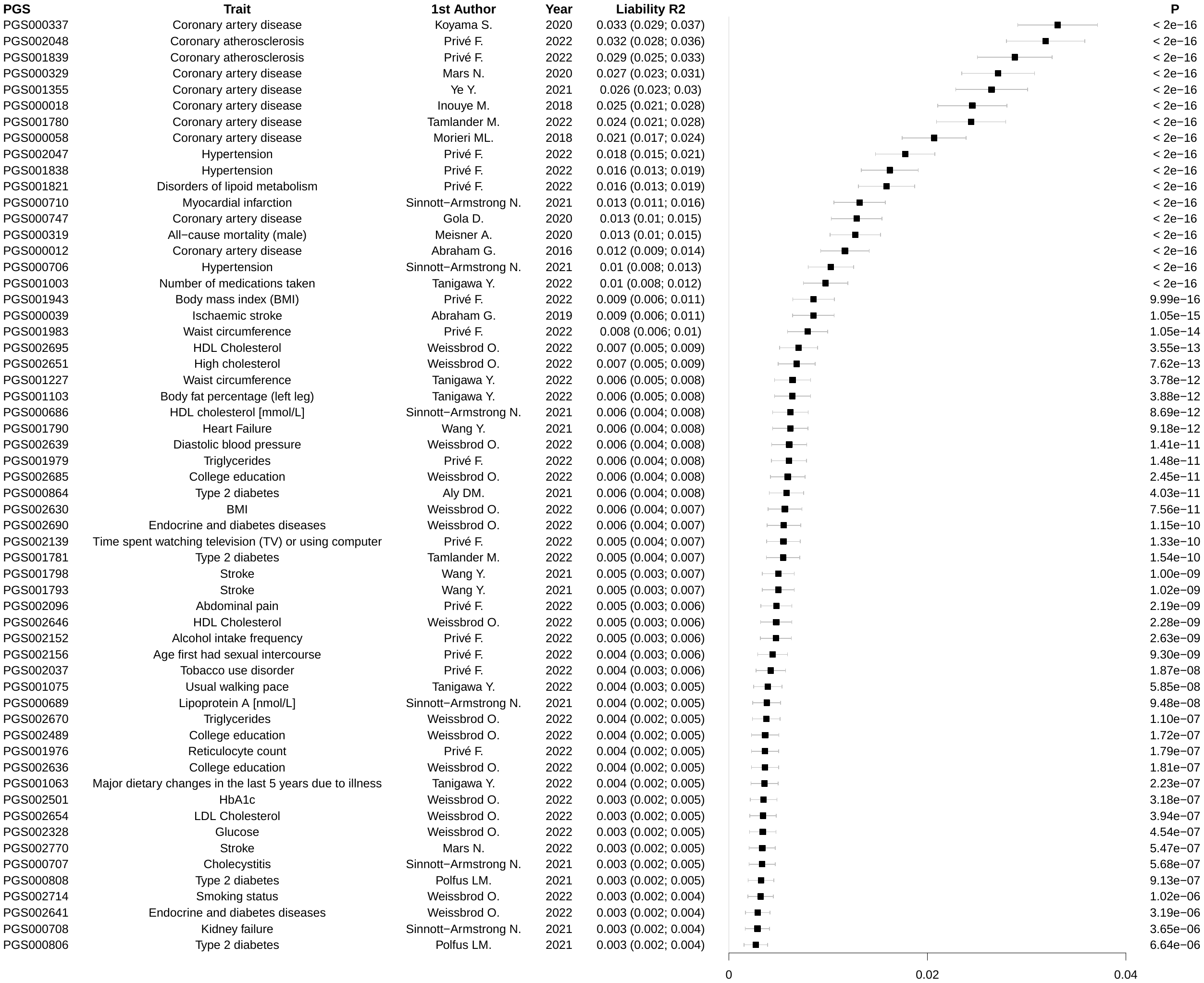

###### **Supplementary Figure 6. Prediction accuracy in the training set of the contributing PGSs to PRSmix+ of coronary artery disease in the South Asian ancestry.**

We used Elastic Net to estimate the mixing weights of multiple PGS and report the contributing PGS with non-zero effects. Prediction accuracy was estimated with liability R2. The boxes represent the mean prediction accuracy across the traits in that group and the whiskers demonstrate 95% confidence intervals.

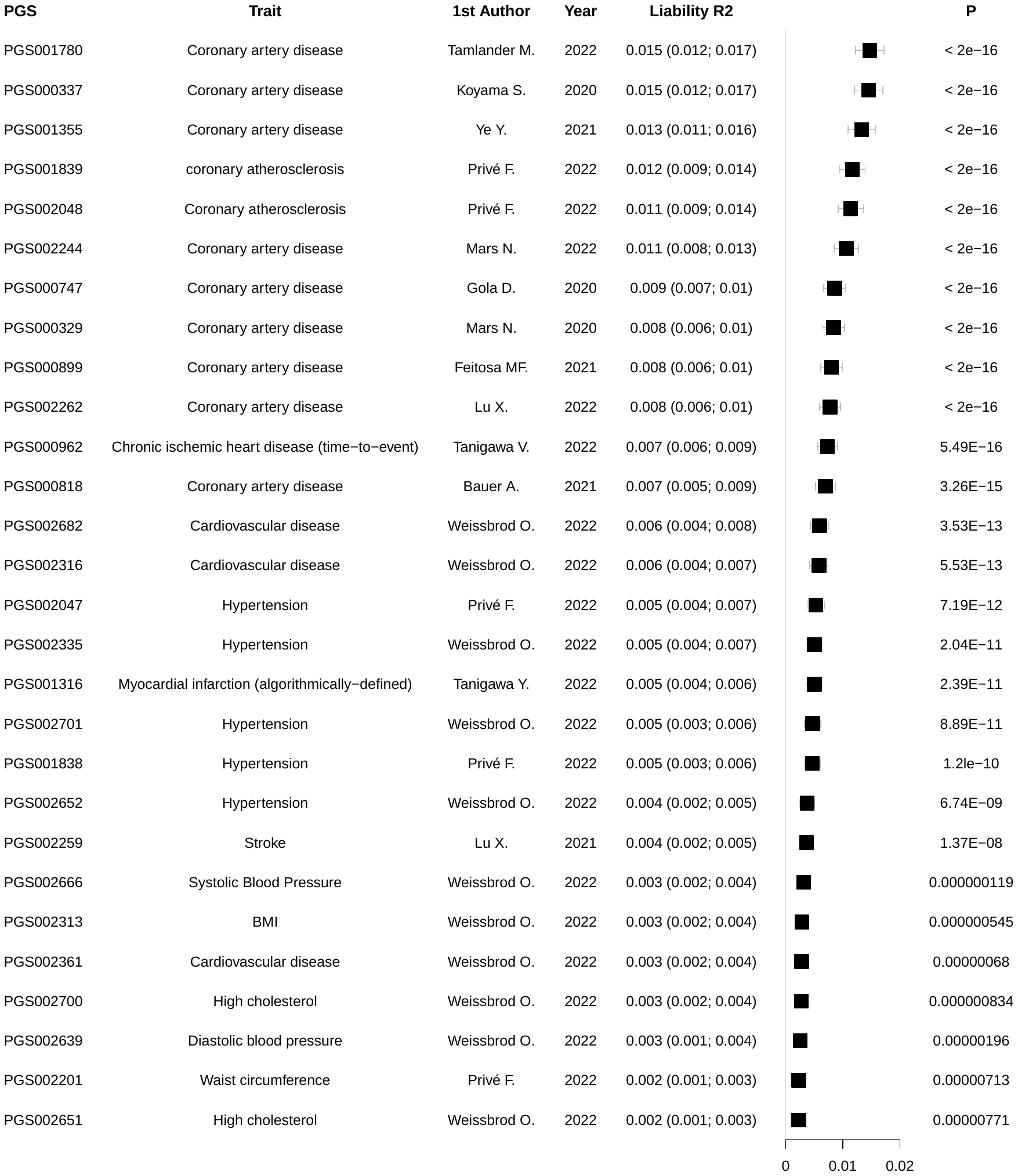

###### **Supplementary Figure 7. Empirical prediction accuracy with various training sample sizes for linear combination for coronary artery disease in the European and South Asian ancestries.**

We assessed the performance of PRSmix and PRSmix+ with various training sample sizes. The prediction accuracy was estimated as liability R2. The points represent the mean prediction accuracy and the whiskers demonstrate 95% confidence intervals.

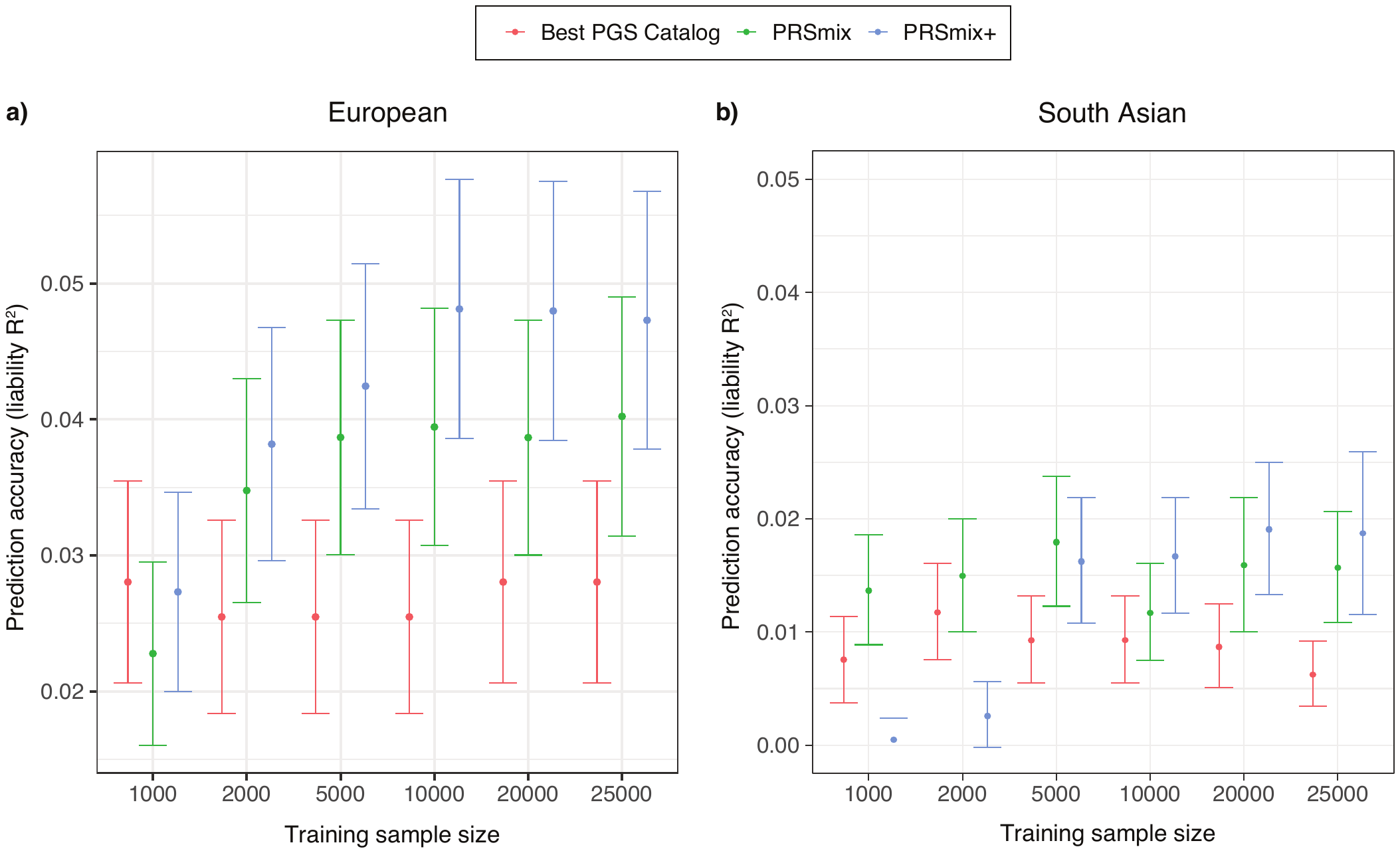

###### **Supplementary Figure 8. Contributing traits to PRSmix+ of stroke in the European ancestry.**

We used Elastic Net to estimate the mixing weights of multiple PGS and report the top 80 contributing PGS with non-zero effects. Prediction accuracy was estimated with liability R2. The boxes represent the mean prediction accuracy across the traits in that group and the whiskers demonstrate 95% confidence intervals.

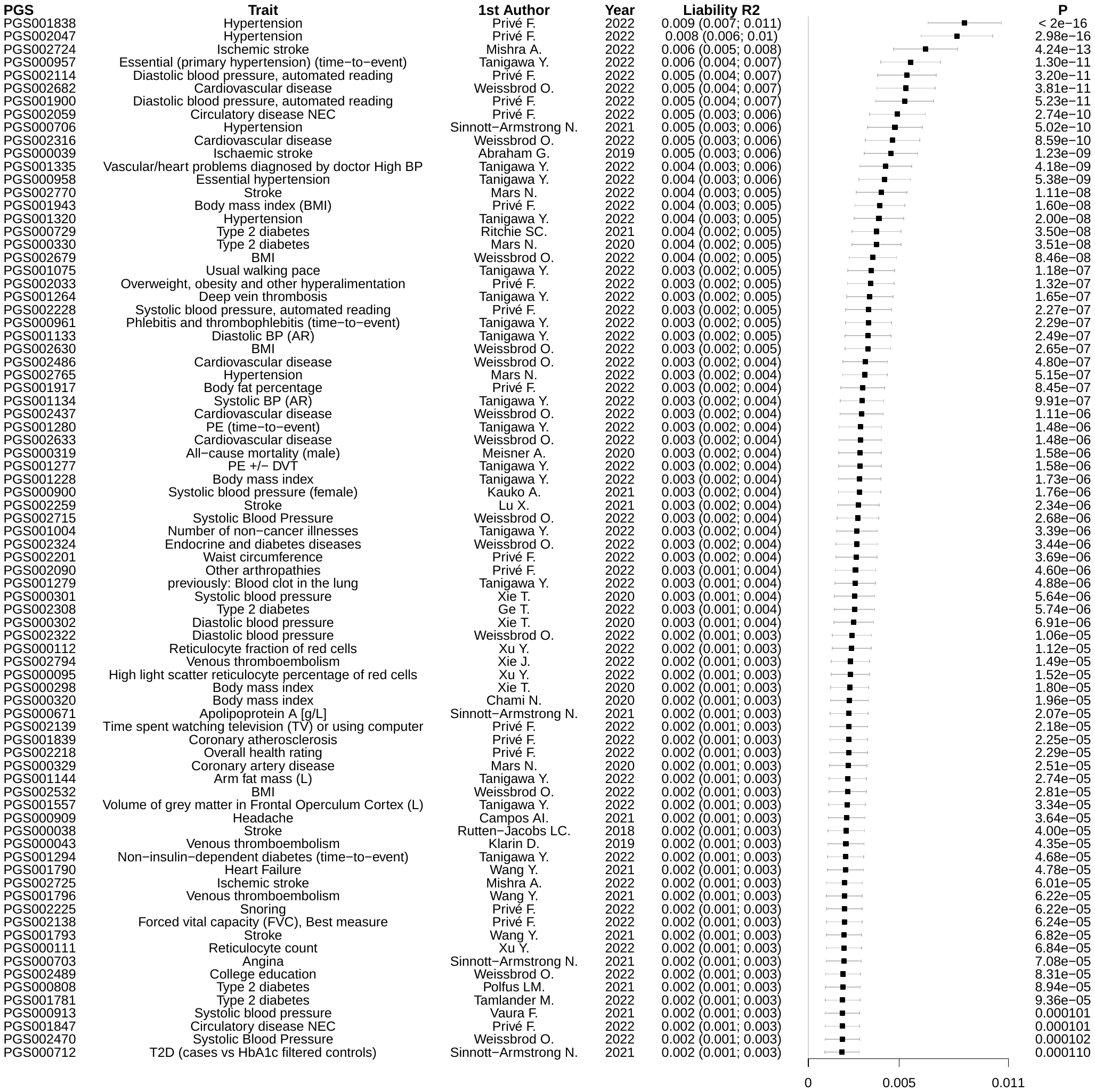

### **Supplementary Table 1. Demographics of European and South Asian ancestries in All of Us and Genes and Health cohorts, respectively.**

ALP, Alkaline Phosphatase; ALT, Alanine transaminase; AST, Aspartate aminotransferase; BMI, Body mass index; Breast CA, Breast cancer; BUN, Blood urea nitrogen; CAD, Coronary artery disease; DBP, Diastolic blood pressure; Hb, Hemoglobin; Hct, Hematocrits; HDL-C, High-density lipoprotein; HTN, Hypertension; LDL-C, Low-density lipoprotein; Prostate CA, Prostate cancer; RA, Rheumatoid arthritis; RBC, Red blood counts; RDW, Red Cell Distribution Width; SBP, Systolic blood pressure; Skin CA, Skin cancer; T2D, Type 2 diabetes; VTE, venous thromboembolism; WBC, White blood counts. NA, not applicable in Genes & Health cohort.

### **Supplementary Table 2. Prediction accuracy of the best PGS catalog, PRSmix and PRSmix+ for 47 traits in the European ancestry from the All of Us cohort.**

The prediction accuracy was assessed as partial R2 and liability R2 for continuous and binary traits, respectively. The partial R2 is a difference of R2 between the model with PRS and covariates including age, sex, and 10 PCs versus the base model with only covariates. Prediction accuracy for binary traits was assessed with liability R2 where disease prevalence was approximately estimated as a proportion of cases in the testing set. ALP, Alkaline Phosphatase; ALT, Alanine transaminase; AST, Aspartate aminotransferase; BMI, Body mass index; Breast CA, Breast cancer; BUN, Blood urea nitrogen; CAD, Coronary artery disease; DBP, Diastolic blood pressure; Hb, Hemoglobin; Hct, Hematocrits; HDL-C, High-density lipoprotein; HTN, Hypertension; LDL-C, Low-density lipoprotein; Prostate CA, Prostate cancer; RA, Rheumatoid arthritis; RBC, Red blood counts; RDW, Red Cell Distribution Width; SBP, Systolic blood pressure; Skin CA, Skin cancer; T2D, Type 2 diabetes; VTE, venous thromboembolism; WBC, White blood counts. NA, not applicable in Genes & Health cohort.

### **Supplementary Table 3. Prediction accuracy of the best PGS catalog, PRSmix and PRSmix+ for 32 traits in the South Asian ancestry from the Genes and Health cohort.**

The prediction accuracies for 32 traits were assessed as partial R2 and liability R2 for continuous and binary traits, respectively. Binary traits with disease prevalence > 2% were selected. The partial R2 is a difference of R2 between the model with PRS and covariates including age, sex, and 10 PCs versus the base model with only covariates. Prediction accuracy for binary traits was assessed with liability-R2 where disease prevalence was approximately estimated as a proportion of cases in the testing set. ALP, Alkaline Phosphatase; ALT, Alanine transaminase; AST, Aspartate aminotransferase; BMI, Body mass index; Breast CA, Breast cancer; BUN, Blood urea nitrogen; CAD, Coronary artery disease; DBP, Diastolic blood pressure; Hb, Hemoglobin; Hct, Hematocrits; HDL-C, High-density lipoprotein; HTN, Hypertension; LDL-C, Low-density lipoprotein; Prostate CA, Prostate cancer; RA, Rheumatoid arthritis; RBC, Red blood counts; RDW, Red Cell Distribution Width; SBP, Systolic blood pressure; Skin CA, Skin cancer; T2D, Type 2 diabetes; VTE, venous thromboembolism; WBC, White blood counts. NA, not applicable in Genes & Health cohort.

### **Supplementary Table 4. Comparisons of PRSmix and PRSmix+ to the best PGS Catalog across groups of traits.**

We categorized all of 47 traits from European ancestry and 31 traits from South Asian ancestry into 6 main groups including anthropometric, blood counts, cancer, cardiometabolic, biochemistry and other conditions. Cancer phenotypes in South Asian ancestry are omitted due to prevalence < 2%.

### **Supplementary Table 5. Benchmarking combination methods with PRSmix and PRSmix+.**

LDpred2-auto was used as the baseline method to input in the methods. 5 traits from Maier et al.^19^ and 26 publicly available GWAS for European ancestry were curated. The components of each combination method are denoted in parentheses. wMT-SBLUP was conducted with the input of sample sizes from the GWAS summary statistics and heritabilities and genetic correlation between all pairs of traits using LD score regression. PRSmix (LDpred2 + PGS Catalog) combined target trait-specific scores within 26 scores and PGS Catalog. Elastic Net (LDpred2) was performed using Elastic Net with all scores from 26 traits generated with LDpred2-auto. PRSmix+ (LDpred2 + PGS Catalog) was conducted using 26 scores from LDpred2-auto and scores from all traits obtained from PGS Catalog. Partial R2 and liability R2 were used for continuous traits and binary traits, respectively. The whiskers demonstrate 95% confidence intervals of mean prediction accuracy. BMI, Body mass index; CAD, coronary artery disease; T2D, type 2 diabetes. GWAS, genome-wide association study.

### **Supplementary Table 6. Ratio of comparison across different methods**

The ratio is estimated as the prediction accuracy of the first method against the second method. 95% confidence intervals were obtained through sub-sampling. P-values were adjusted for false discovery rate.

### **Supplementary Table 7. Net reclassification improvement of the best PGS Catalog, PRSmix and PRSmix+ compared to the baseline model with clinical risk factors.**

The baseline model for risk prediction includes age, sex, total cholesterol, HDL-C, systolic blood pressure, BMI, type 2 diabetes and current smoking status. The clinical risk factors includes age, sex, EUR, European; SAS, South Asian. NRI, Net Reclassification improvement.

### **Supplementary Table 8. Incremental AUC for CAD PRSs in European and South Asian ancestries.**

The incremental AUC was estimated as the difference between AUC of the full model of PRS and covariates including age, sex and 10 PCs, and the null model with only covariates. The risk was adjusted for age, sex and 10 PCs. EUR, European; SAS, South Asian.

### **Supplementary Table 9. Risk stratification for CAD with PRSs in European and South Asian ancestries.**

Odds ratios were calculated by comparing those with PRS in the top decile versus the remaining of population in a logistic regression in each ancestry. The risk was adjusted for age, sex and 10 PCs. EUR, European; SAS, South Asian.

### **Supplementary Table 10. PheWAS analysis of the best PRS estimated from PGS Catalog and PRSmix+ with disease phenotypes for European ancestry in the All of Us cohort.**

See the Excel file

###### **Supplementary Table 11. Fold difference of odds ratio between PRSmix+ and the best PGS Catalog for each PheWAS category.**

The fold-difference was calculated as a ratio between odds ratio of PRSmix+ versus odds ratio of the best PGS Catalog. P-value was estimated from a pair-T test across each pair in each pheWAS category.

See the Excel file

### **Supplementary Table 12. Mixing weights of PGS Catalog scores with PRSmix in European ancestry**

See the Excel file

### **Supplementary Table 13. Mixing weights of PGS Catalog scores with PRSmix+ in European ancestry**

See the Excel file

### **Supplementary Table 14. Mixing weights of PGS Catalog scores with PRSmix in South Asian ancestry**

See the Excel file

### **Supplementary Table 15. Mixing weights of PGS Catalog scores with PRSmix+ in South Asian ancestry**

See the Excel file

### **Supplementary Table 16. Phenotype Definitions and Descriptions from the All of Us Research Program.**

For binary phenotypes, cases were curated as per the definitions below. Controls were defined as those who did not meet the case definition, and who had consented to linking of EHR data. 1.9% of participants (6,692/313,514 (1.9%)) of participants did not consent to EHR data linkage in the V6 Data Release.

Continuous measures included either body measurements, which were directly measured at the participant’s study visit, or laboratory measurements, obtained from electronic health record data. For all laboratory measurements, if a participant had multiple measurements available, the most recent measurements was used (closest to CDR date) after cleaning.

### **Supplementary Table 17. Phenotype Definitions and Descriptions from the Genes and Health cohort.**

For binary phenotypes, cases were curated as per the definitions below. Controls were defined as those who did not meet the case definition, and who had consented to linking of EHR data.

Continuous measures included either body measurements, which were directly measured at the participant’s study visit, or laboratory measurements, obtained from electronic health record data. For all laboratory measurements, if a participant had multiple measurements available, the average was used after cleaning.

### **Supplementary Table 18. Publicly available summary statistics.**

ADHD, attention deficit hyperreactive disorder; BMI, body mass index; CAD, coronary artery disease; RA, rheumatoid arthritis; TG, triglycerides; T2D, type 2 diabetes; WHR, waist-hip-ratio.
